# Reporting Quality in Health Economic Evaluation Studies of Immune Checkpoint Inhibitors: A Systematic Review

**DOI:** 10.1101/2024.01.24.24301756

**Authors:** Takashi Yoshioka, Shintaro Azuma, Satoshi Funada, Takahiro Itaya, Rei Goto

## Abstract

**Objectives:** This study assessed the reporting quality of health economic evaluation (HEE) studies of immune checkpoint inhibitors (ICIs).

**Methods:** We conducted a systematic literature search of four databases (PubMed, EMBASE, Cochrane CENTRAL, and the International HTA Database) for studies published between January 1, 2014 and December 31, 2022. Three pairs of reviewers independently screened and reviewed the full text and extracted the data. We included all ICIs approved up to December 31, 2022, in the United States (US), European Union, China, and Japan. Reporting quality was assessed using the Consolidated Health Economic Evaluation Reporting Standards published in 2013 (CHEERS 2013). Subgroup analyses were also performed based on the risk of sponsorship bias or citation of CHEERS 2013.

**Results:** A total of 5,368 records were identified, 252 of which were included after full-text review. The study design, setting, and ICIs most frequently observed were cost-effectiveness and cost-utility analyses (63.5%), the US (46.0%), and pembrolizumab (38.1%), respectively. Of the 24 items of CHEERS 2013, fully reported items were limited, particularly in the Methods section. Setting and location were not reported in 94.4% of the records. Similar trends were observed in subgroup analysis.

**Conclusion:** HEE studies on ICIs between 2014 and 2022 had limited reporting across the 24 items of CHEERS 2013, regardless of sponsorship bias risk or citations. The items on setting and location in the Methods section were particularly underreported, emphasizing the need for transparent reporting in HEE studies of ICIs.

**Highlights:**

- The reporting quality of health economic evaluation (HEE) studies was evaluated using the Consolidated Health Economic Evaluation Reporting Standards published in 2013 (CHEERS 2013). However, the reporting quality of HEEs of immune checkpoint inhibitors (ICIs), which is an emerging health policy issue for the economic burden of cancer, remains unknown.
- Despite the passage of a decade since the publication of CHEERS 2013, HEE studies on ICIs have generally not fully adhered to the CHEERS 2013 guidelines in the Methods section. This is particularly evident in the setting and location items, even after stratification by the presence or absence of risk of sponsorship bias or whether CHEERS 2013 statement was cited.
- This study highlights the insufficient reporting of CHEERS items among current HEE studies of ICIs, especially in the Methods section, to researchers who conduct HEE studies of ICIs, and informs policymakers and stakeholders who refer to HEE studies of ICIs about underreporting.

## Introduction

The economic burden of cancer represents one of the most critical issues in the context of health policy worldwide, along with its huge disease burden.^1,2^ It accounts for large healthcare spending for both patients and insurers, and productivity loss due to employment loss, absenteeism, presenteeism, and premature deaths.^2–4^ The economic burden is substantial, as the estimated global cost of cancer from 2020 to 2050 is 25.2 trillion international dollars.^2^ Considering the economic impact of cancer care, based on the wide range of cancer economic burdens and finite resources in healthcare systems, it is crucial for policymakers to develop effective policies to manage the expected increase in cancer prevalence and improve morbidity and mortality.

Among cancer-related economic impacts, an increase in healthcare spending, particularly due to increasing anticancer drug prices, is a major global challenge. Increased anticancer drug prices affect individual treatment access, improvement of cancer-related outcomes, and management of resources in healthcare systems.^5^ Among the anticancer agents, immune checkpoint inhibitors (ICIs), which are the cornerstone of cancer immunotherapy that have undergone remarkable development over the past two decades, have contributed substantially to growing costs.^6,7^ Evidence shows that expenditures of ICIs increased from 2.8 million to 4.1 billion dollars between 2011 and 2021 in the United States Medicaid Programs.^8^ Given such financial impacts, ICIs have led to discussions on the necessity of considering economic burden in addition to clinical efficacy.

Health economic evaluation (HEE) is one way to simultaneously examine economic costs and clinical benefits from the perspective of the health sector and society. Well-designed HEEs with clear principles and rigorous methodology can provide policymakers with evidence-based recommendations to help effective resource allocation.^9^ Therefore, several countries have used HEE-based guidance from third-party agencies, *e.g.*, the National Institute for Health and Care Excellence (NICE) in the United Kingdom (UK), for insurance coverage and/or price adjustment of new health technologies.^10^ Along with HEEs from these agencies, numerous HEE studies have been conducted by researchers and manufacturers to help policymakers in decision-making.^11,12^ To make HEE studies useful for policy decision-making, it is essential to clarify the application of robust methodologies, development of valid decision models with appropriate data and assumptions, and evaluation of uncertainty.^11^ However, fully covering these within the limited pages of journals has been challenging, and the reporting of HEE research has long been an issue from the perspective of reporting quality improvement.^13^

To address this, the International Society for Pharmacoeconomics and Outcomes Research (ISPOR) developed and published reporting standards, *i.e.*, the Consolidated Health Economic Evaluation Reporting Standards (CHEERS).^14^ The CHEERS checklist, first published in 2013 and revised in 2022,^14,15^ is available through the Enhancing the Quality and Transparency Of Health Research (EQUATOR) network.^16^ It has been translated into several languages and is referenced in multiple HEE studies worldwide.^17^ To date, the CHEERS statement has been reported in several systematic reviews of HEE studies or in those focusing on the quality of reporting in HEE research.^18–22^ However, most studies have used it as a scoring tool, which is stated in the standards as misuse of the tool,^15^ or in the context of assessing methodological quality, resulting in few accurate assessments of reporting transparency. Furthermore, there is only one previous systematic review of HEEs of ICIs.^23^ However, the review documented HEEs of specific ICIs available until April 1, 2018, and did not assess the quality of reporting.

Owing to the increasing importance of HEEs for ICIs, there is a rising demand from both academic and policy sectors to systematically and appropriately assess the quality of reporting in HEE studies. In this context, this study aimed to systematically review the HEEs of all approved ICIs through 2022 and to assess the quality of reporting using CHEERS.

## Methods

### Study Design

We conducted a systematic review in accordance with the Preferred Reporting Items for Systematic Reviews and Meta-Analyses (PRISMA) standards.^24^ We registered the protocol for this study in PROSPERO (CRD42023439699).

### Inclusion and exclusion criteria

We included cost-effectiveness, cost-utility, cost-benefit, and cost-minimization studies for the HEEs of ICIs. We focused on all neoplasms eligible for ICIs, including hematological diseases and sarcomas. Eligible ICIs were based on approvals through December 31, 2022, by the United States Food and Drug Administration (FDA), the European Medicines Agency (EMA), the Pharmaceuticals and Medical Devices Agency (PMDA) of Japan, and the National Medical Products Administration (NMPA) of China.^25^ The included ICIs were as follows: atezolizumab, avelumab, cadonilimab, camrelizumab, cemiplimab, dostarlimab, durvalumab, envafolimab, ipilimumab, nivolumab, pembrolizumab, penpulimab, relatlimab, retifanlimab, serplulimab, sintilimab, sugemalimab, tislelizumab, toripalimab, tremelimumab, and zimberelimab. As this study focused on the HEEs of ICIs approved through December 31, 2022, we assumed that CHEERS 2022, published in January 2022, had not yet been adequately disseminated to investigators at the time the HEEs of these ICIs were conducted. Therefore, we excluded studies that cited the CHEERS 2022 statement. Publication type was limited to original articles, and language was limited to English.^26^

### Search Strategy

We extracted all studies from four databases, all of which have been discussed and used by researchers and librarians: PubMed, EMBASE, Cochrane CENTRAL, and the International HTA database. As noted above, this study focused on CHEERS 2013, and we assumed that it took several months for researchers to adopt CHEERS 2013, published in March 2013, as a reporting standard for HEE studies. Therefore, this study defined the search period as January 1, 2014, to December 31, 2022. The search strategies were constructed by two librarians at the authors’ institute, and actual searches were conducted on July 7, 2023. The search terms are listed in **Appendix Table 1**.

**Table 1.** Baseline characteristics.

| **Characteristics** | **Overall**<br>**n = 252** |
| --- | --- |
| **Study design, *n* (%)** |  |
| Cost-effectiveness and cost-utility analyses | 160 (63.5) |
| Cost-utility analysis | 69 (27.4) |
| Cost-effectiveness, cost-utility, and cost-benefit analyses | 13 (5.2) |
| Cost-effectiveness analysis | 5 (2.0) |
| Others | 5 (2.0) |
| **Setting, *n* (%)** |  |
| US | 116 (46.0) |
| China | 66 (26.2) |
| US and China | 12 (4.8) |
| Others | 50 (19.8) |
| Not reported | 8 (3.2) |
| **Target population, *n* (%)** |  |
| Non-small cell lung cancer | 83 (32.9) |
| Melanoma | 31 (12.3) |
| Renal cell carcinoma | 28 (11.1) |
| Esophageal cancer | 20 (7.9) |
| Others | 90 (35.7) |
| **Intervention<sup>\*</sup>, *n* (%)** |  |
| Pembrolizumab | 96 (38.1) |
| Nivolumab | 44 (17.5) |
| Atezolizumab | 36 (14.3) |
| Ipilimumab + Nivolumab | 30 (11.9) |
| Others | 71 (28.2) |
| **Study perspective, *n* (%)** |  |
| Health system or payer perspective | 219 (86.9) |
| Societal perspective | 12 (4.8) |
| Others | 13 (5.2) |
| Not reported | 8 (3.2) |
| **Time horizon, *n* (%)** |  |
| Lifetime | 90 (35.7) |
| Non-lifetime | 148 (58.7) |
| Not reported | 14 (5.6) |
| **Risk of sponsorship bias, *n* (%)** |  |
| Present | 84 (33.3) |
| Absent | 168 (66.7) |
| **Citation of the CHEERS<sup>1</sup> statement, *n* (%)** |  |
| Cited | 37 (14.7) |
| Not cited | 215 (85.3) |

Notes. CHEERS 2013, the Consolidated Health Economic Evaluation Reporting Standards published in 2013, United States. \* Some of the included reports conducted health economic evaluations of multiple checkpoint inhibitors within multiple arms. Consequently, the total number of interventions exceeded the total sample size (n=252), resulting in the sum of the percentages exceeding 100%.

### Study Selection and Data Extraction

After excluding duplicate records, three independent pairs of researchers (SA and TY, SA and SF, and SA and TI) screened the titles and abstracts of the articles found in the literature search. The same pairs of researchers independently screened the full text of each study. After screening the full-text records, the same independent pairs of researchers used pre-specified data extraction forms to collect data from the included HEE studies. Any disagreements were resolved through consensus discussions.

The extracted data included general information and the quality of reporting. General information included study design, setting, target population, intervention (*i.e.*, ICIs), study perspective, time horizon, publication year, risk of sponsorship bias, and citation of CHEERS 2013. The risk of sponsorship bias was defined as present if the authors of the study included employees of the manufacturer or if they received funding from the manufacturer. The quality of reporting will be described independently later.

### Outcome (Quality of Reporting)

The outcome of interest was the reporting quality. The quality of reporting among the included studies was evaluated using the CHEERS 2013 checklist.^14^ CHEERS 2013 includes 24 items divided into six main categories to provide systematic reporting of HEEs. With reference to CHEERS 2013, several items required reporting of multiple aspects (*e.g.*, item 2: “*Provide a structured summary of the objectives, perspective, setting, methods [including study design and inputs], results [including base case and uncertainty analyses], and conclusions.*”). To distinguish between full and partial reporting of these components, we developed a unique checklist based on CHEERS 2013 (**Appendix Table 2**). Studies were assessed as “fully reported” if they met all checklist items, “partially reported” if only some were met, “not reported” if none were met, and “not applicable” if irrelevant.

### Statistical Analysis

First, the characteristics of the included studies were summarized as numbers and proportions (%) of categorical variables. We also described the annual publication numbers of the studies, distinguishing them based on whether the CHEERS statement was cited. We then described the quality of reporting results by presenting the numbers of the categorical outcome variables (“fully reported,” “partially reported,” and “not reported”) for each of the 24 items. We assumed that the quality of reporting might differ by the presence of sponsorship bias.^27^ Furthermore, we assumed that a declaration to follow CHEERS 2013 and citing the statement may influence the researcher’s attitude toward transparent reporting. Given this, we performed subgroup analyses that presented the proportions of “fully reported” outcomes among all outcome variables excluding “not applicable” for each item by the presence of “risk of sponsorship bias” or “citation of CHEERS 2013.”

### Ethical consideration

This study was deemed exempt from review by the institutional review board based on the ethical guidelines owing to the study design (systematic review).

## Results

A flow diagram of the study is shown in **Figure 1**. We identified 5,368 records, 344 of which were eligible after screening the titles and abstracts. Of the 344 records, we finally included 252 were eligible after full-text screening. A summary of the included studies is presented in **Appendix Table 3**.

**Figure 1.**
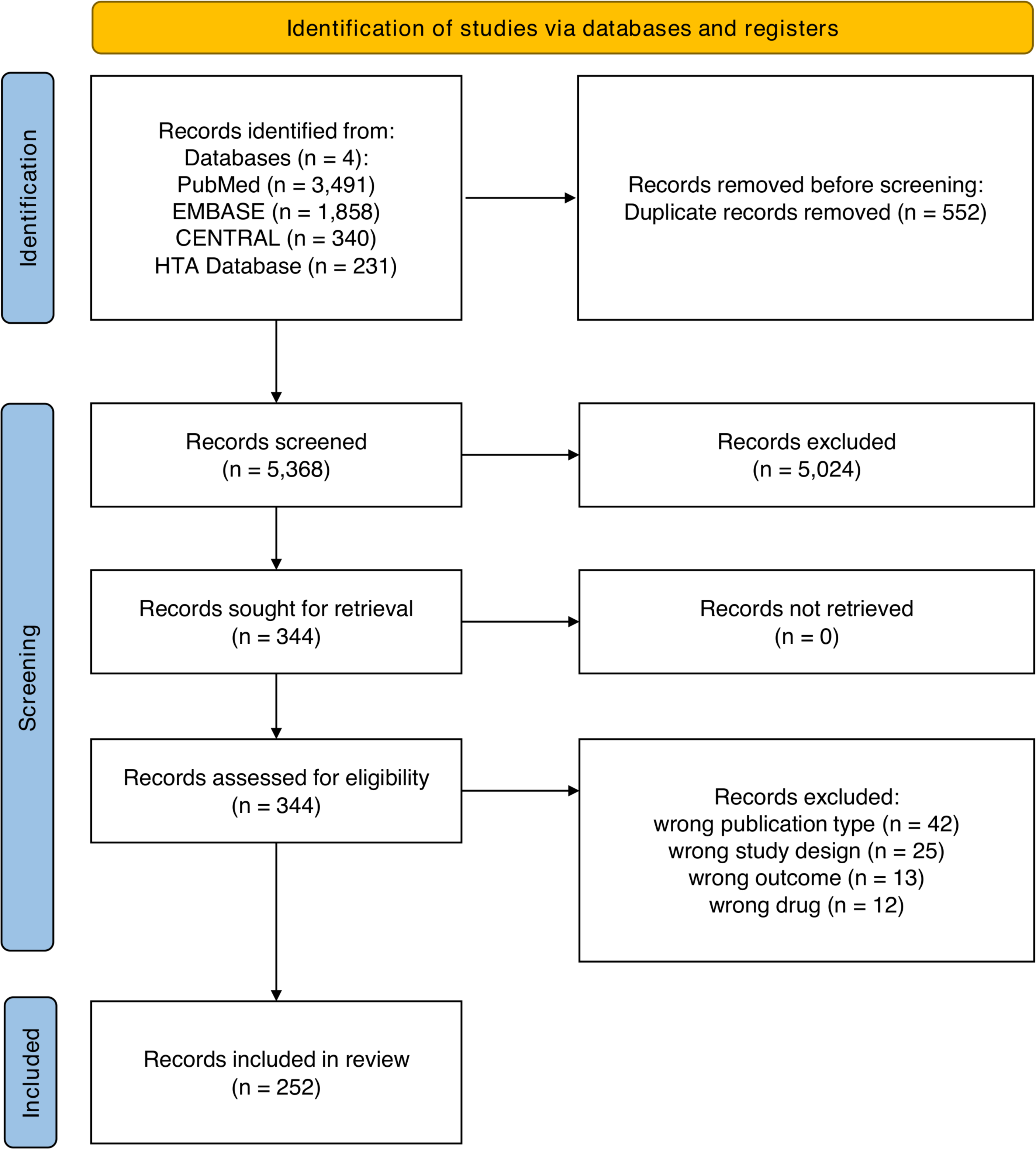
PRISMA 2020 flow diagram. Notes. PRISMA 2020, the Preferred Reporting Items for Systematic reviews and Meta-Analyses updated in 2020.

**Table 1** shows the baseline characteristics of the records. Of the 252 records, 160 (63.5%) were subjected to the cost-effectiveness and cost-utility analyses. The settings were mostly in the United States (n=116, 46.0%) and China (n=66, 26.2%). Non-small cell lung cancer (n=83, 32.9%), melanoma (n=31, 12.3%), and renal cell carcinoma (n=28, 11.1%) were also observed. The most common interventions (*i.e.*, ICIs) were pembrolizumab (n=96, 38.1%), nivolumab (n=44, 17.5%), and atezolizumab (n=36, 14.3%). Most studies were conducted from a health system or payer’s perspective (n=219, 86.9%) and non-lifetime horizons (n=148, 58.7%). Eighty-four studies (33.3%) were at risk of sponsorship bias, and only 37 (14.7%) cited CHEERS 2013. As shown in **Figure 2**, the number of publications showed an annual increase, and the proportion of CHEERS 2013 citations showed a substantial increase from 2021.

**Figure 2.**
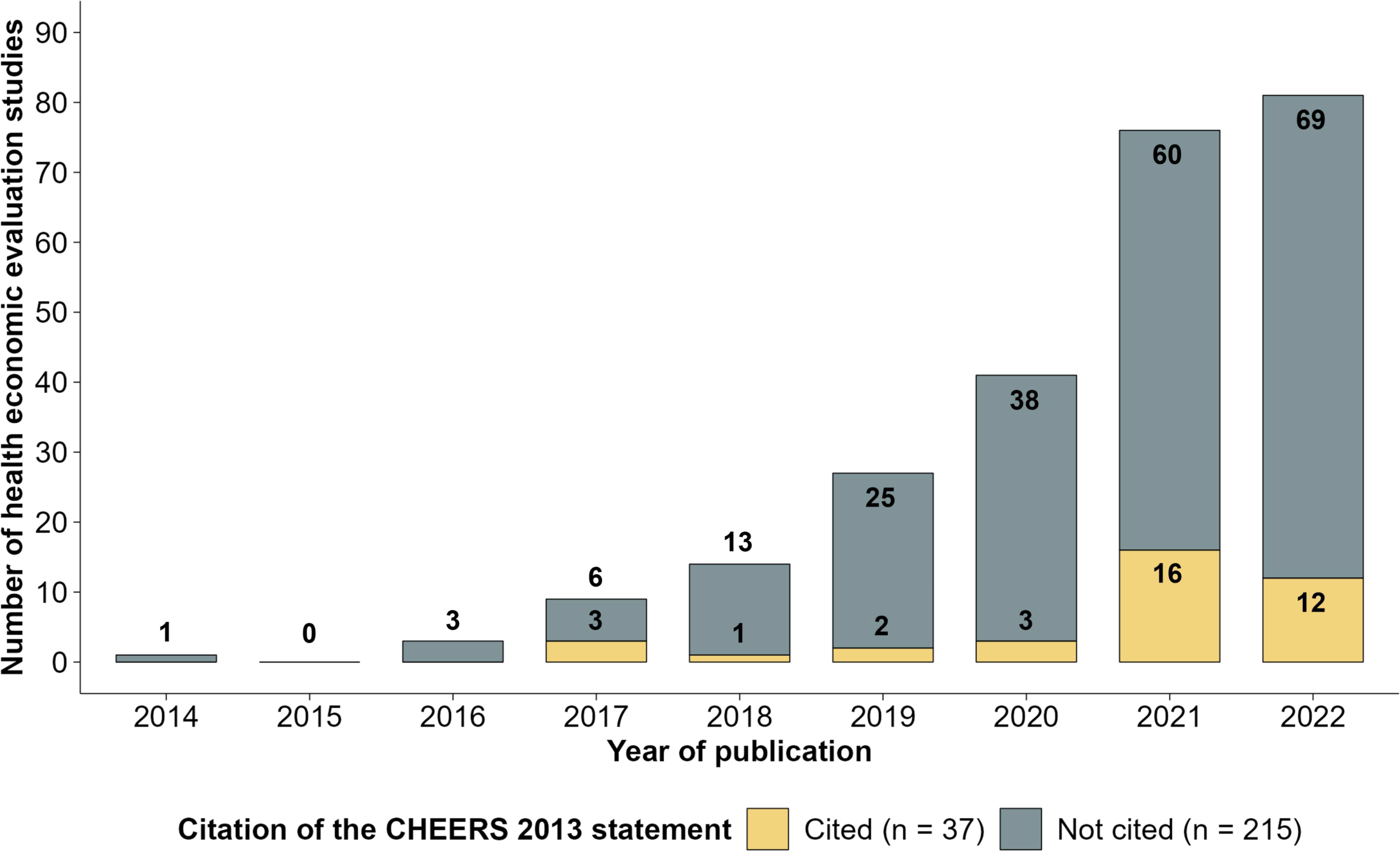
Number of health economic evaluations stratified by whether CHEERS 2013 was cited. Notes. CHEERS 2013, the Consolidated Health Economic Evaluation Reporting Standards published in 2013.

The overall quality of reporting for the study is shown in **Figure 3**. Of the 24 items, the most “fully reported” items were frequently observed for the “Title and abstract,” “Results,” and “Other” (except “source of funding”) sections (*e.g.*, title [n=244, 96.8%], estimating resources and costs [n=237, 94.0%], study parameters [n=231, 91.7%], incremental costs and outcomes [n=248, 98.4%], characterizing uncertainty [n=235, 93.3%], and conflicts of interest [n=249, 98.8%]). In contrast, the least “fully reported” items were observed in the “Methods” section (*e.g.*, setting and location [n=14, 5.6%], study perspective [n=94, 37.3%], choice of health outcomes [n=75, 29.8%], measurement of effectiveness [n=80, 31.7%], analytic methods [n=55, 21.8%], and source of funding [n=87, 34.5%]). Of the least “fully reported” items, setting and location were predominantly “not reported” (n=238, 94.4%).

**Figure 3.**
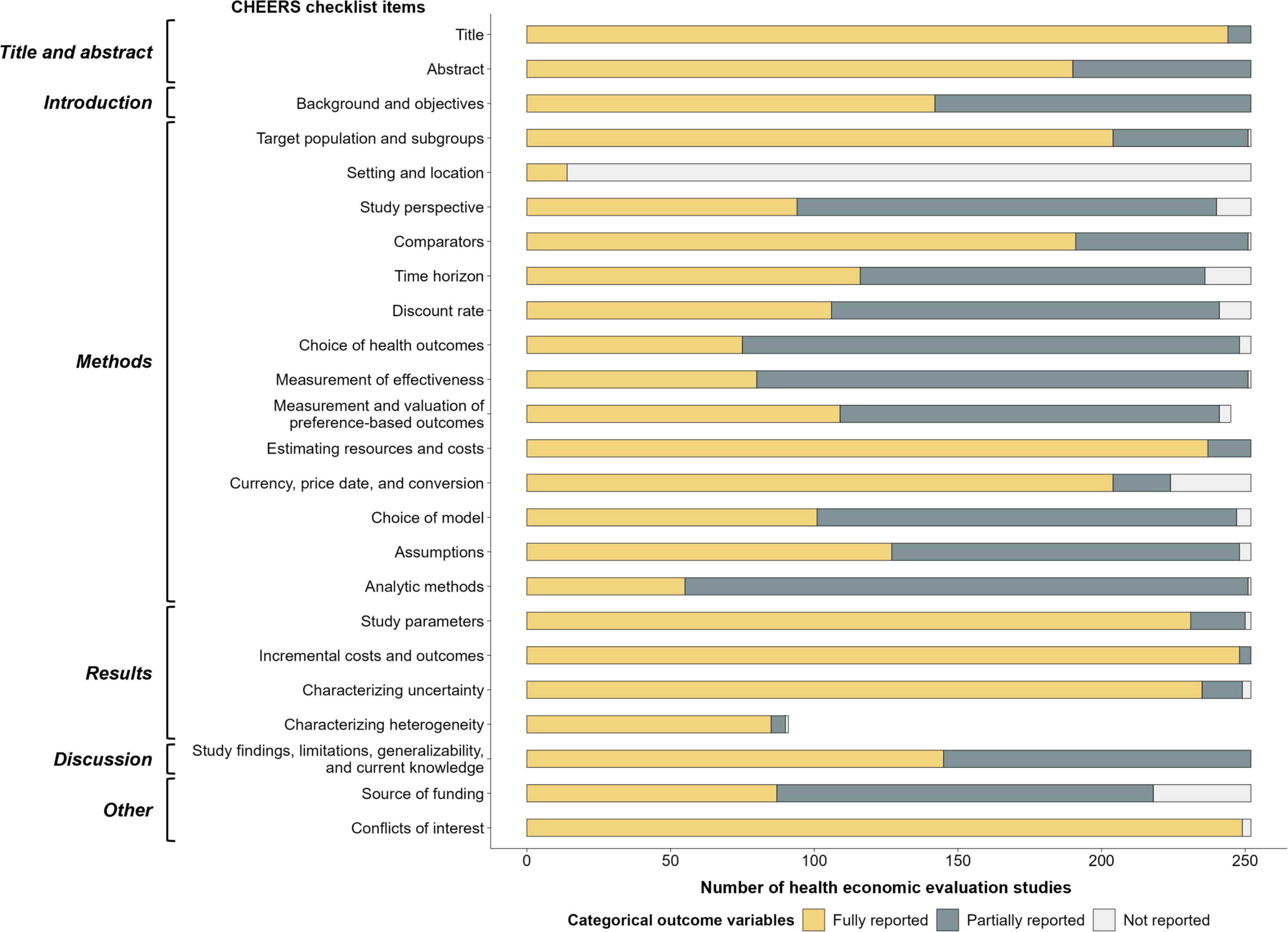
Reporting quality of health economic evaluation studies according to the CHEERS 2013 checklist. Notes. CHEERS 2013, the Consolidated Health Economic Evaluation Reporting Standards published in 2013.

The results of subgroup analyses are shown in **Figure 4** and **Figure 5**. Similar to the results for the overall studies, the most “fully reported” items were observed in the “Title and abstract,” “Results,” and “Other” (except “source of funding”) sections, whereas the least “fully reported” items were observed in the Methods section for both subgroup analyses. HEE studies at the risk of sponsorship bias were more likely to report the measurement and valuation of preference-based outcomes (present, 75.9% vs. absent, 29.5%) and assumptions (present, 70.2% vs. absent, 40.5%), whereas they were unlikely to report the choice of health outcomes (present, 15.5% vs. absent, 36.9%). Interestingly, studies with a risk of sponsorship bias had a higher proportion of “fully reported” for the “source of funding” item compared to those without, although both proportions were low (present, 38.1% vs. absent, 32.7%) (**Figure 4**). Focusing on the citations of CHEERS 2013, HEE studies with citations were more likely to report many of the CHEERS 2013 items than those without. Among the items, the choice of health outcomes (cited, 51.4% vs. not cited, 26.0%) and the measurement of effectiveness (cited, 43.2% vs. not cited, 29.8%) adhered well (**Figure 5**).

**Figure 4.**
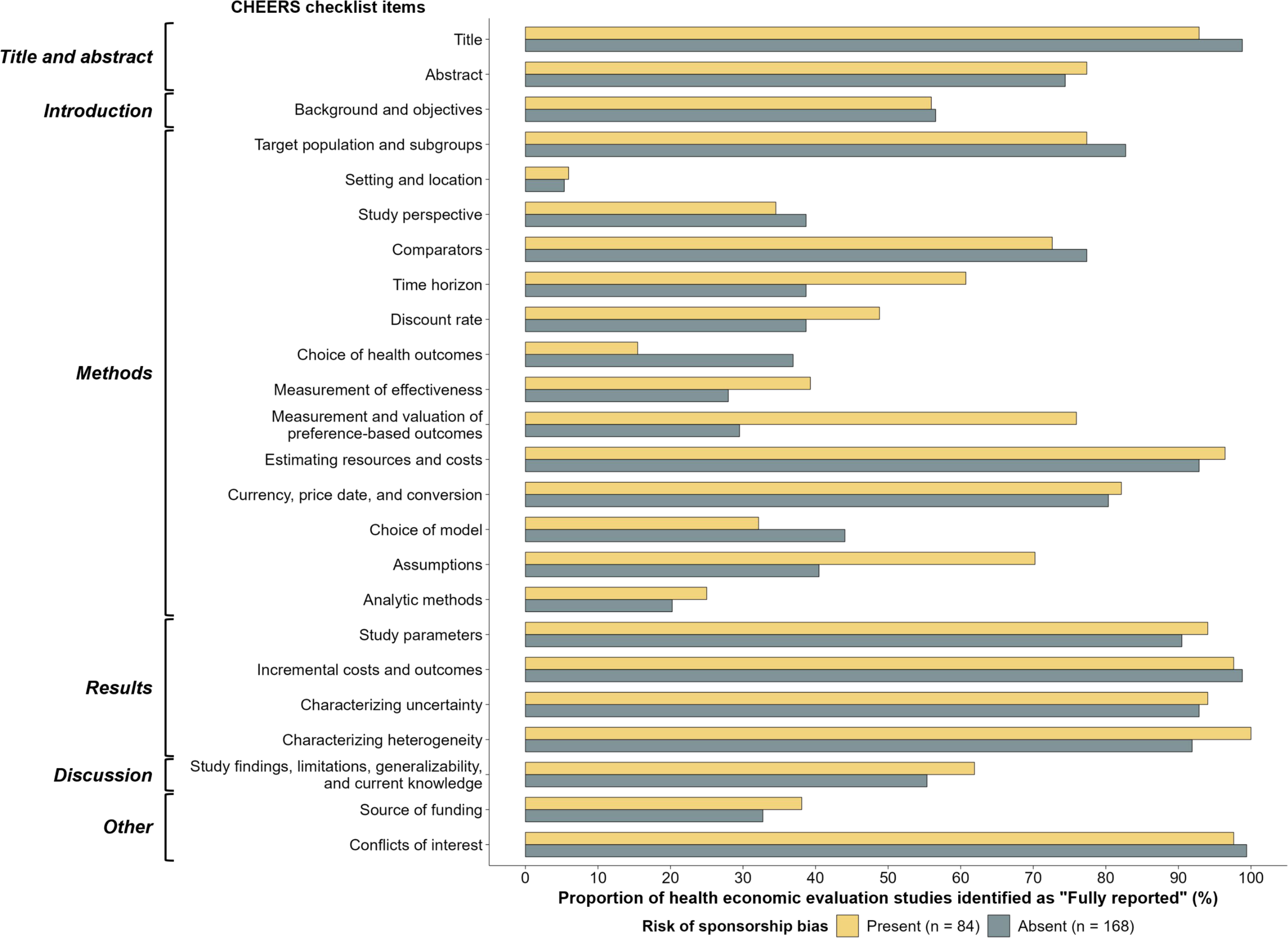
Proportions of the “fully reported” category classified into the presence or absence of the risk of sponsorship bias. Notes. CHEERS 2013, the Consolidated Health Economic Evaluation Reporting Standards published in 2013. In the “measurement and valuation of preference-based outcomes” item, five studies were excluded from the “present” group, and two studies were excluded from the “absent” group; and in the “characterizing heterogeneity” item, 67 studies were excluded from the “present” group and 94 studies were excluded from the “absent” group.

**Figure 5.**
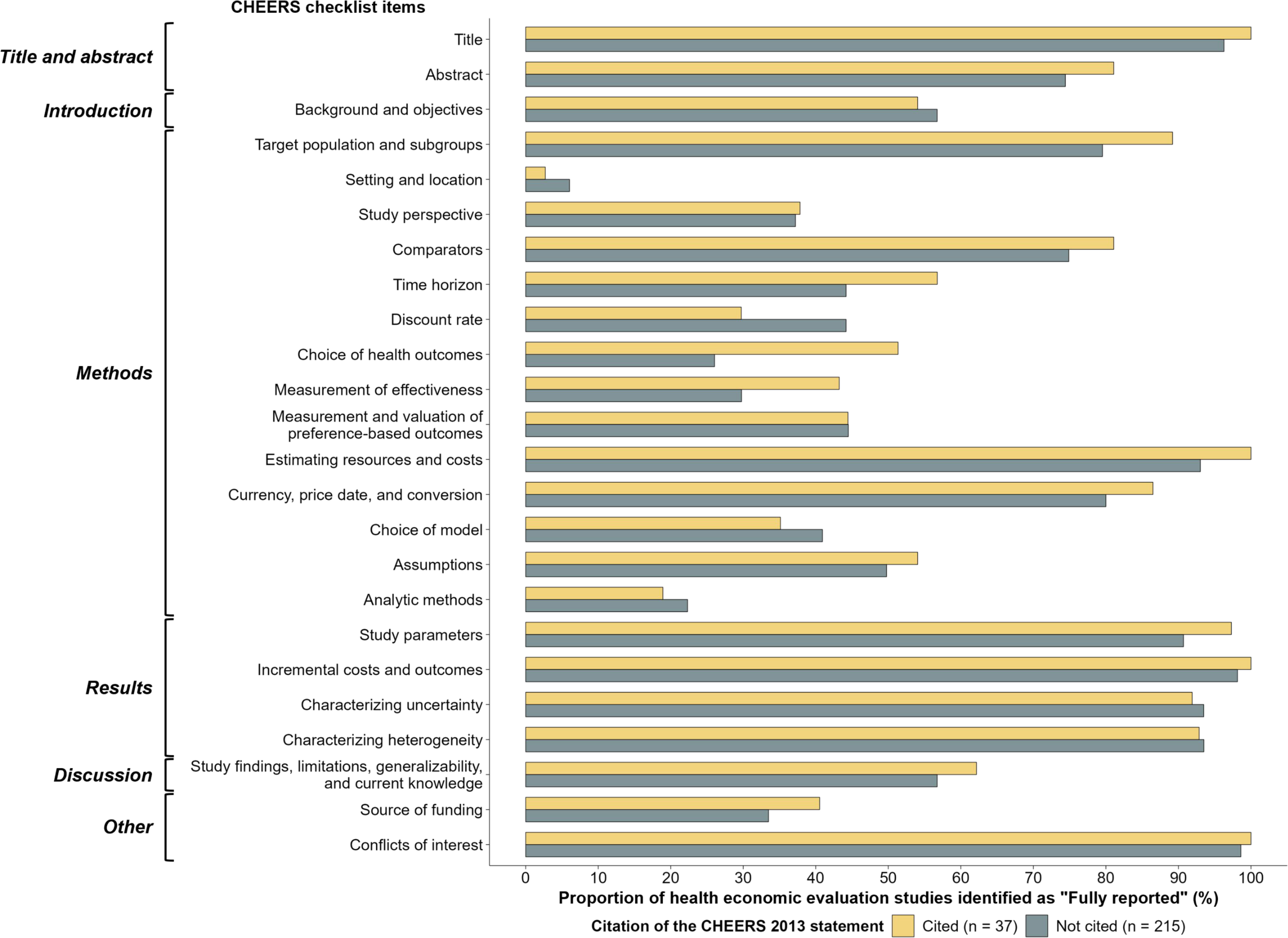
Proportions of the “fully reported” category classified into whether citing the CHEERS 2013 statement or not. Notes. CHEERS 2013, the Consolidated Health Economic Evaluation Reporting Standards published in 2013. In the “measurement and valuation of preference-based outcomes” item, a study was excluded from the “cited” group and six studies were excluded from the “not cited” group; and in the “characterizing heterogeneity” item, 23 studies were excluded from the “cited” group and 138 studies were excluded from the “not cited” group.

## Discussion

In this systematic review, 252 HEE studies on ICIs were conducted between 2014 and 2022. The number of published studies has shown an annual increase, and the number of citations of CHEERS 2013 showed a clear increase since 2021. Of the 24 items of CHEERS 2013, substantially “fully reported” items were limited, observed only for the “Title and abstract,” “Results,” and “Other” (except “source of funding”) sections. The least “fully reported” items were observed in the “Methods” section, and the item of setting and location was poorly adhered to. Similar reporting trends were observed even after stratification by the risk of sponsorship bias or citation of CHEERS 2013. In the analyses based on the presence or absence of sponsorship bias risk, some variations in adherence to specific items in CHEERS 2013 were observed. However, studies that cited CHEERS 2013 generally had high adherence to most items compared with studies that did not cite this statement.

Several characteristics were observed in this study, and the potential mechanisms were considered for each. First, many of the included studies used both cost-effectiveness and cost-utility designs from a health system or payer’s perspective, and were from the United States or China, where the number of patients eligible for ICIs is large.^2^ Such demographic characteristics imply that researchers or manufacturers may provide information on resource allocation for policymakers. Second, the number of studies citing CHEERS 2013 remained limited, even though the number of HEE studies on ICIs has increased annually. The CHEERS 2013 statement may not be well received by researchers conducting HEE studies on ICIs. Evidence indicates that more than 80% of PubMed-indexed pharmacoeconomic studies published between 2021 and 2022 did not declare adherence to the CHEERS statement.^28^ Third, several of the methodological items were only partially reported. In particular, the settings and locations were often not reported at all. This may indicate that the country adjustment emphasized by Drummond et al.,^13^ which is necessary for discussing the transferability of results,^29^ may not be valued sufficiently by researchers. Fourth, even in the presence of the risk of sponsorship bias, there were several items for which the transparency of methodological reporting was high. This may be influenced by the fact that manufacturers conduct HEEs evaluated by third-party agencies, such as NICE in the UK,^30^ in parallel with or prior to HEE studies. Fifth, although not statistically tested, studies citing the CHEERS 2013 statement were found to have more “fully reported” items than those that did not. Citing CHEERS 2013 may contribute to careful reference to and adherence to this statement.

To date, several systematic reviews have evaluated the quality of reporting using the CHEERS 2013 statement. Tai *et al*. used CHEERS 2013 in their systematic review of HEEs conducted from a patient’s perspective.^18^ In their study, CHEERS 2013 was evaluated using four categories for each item: ’’fully satisfied (FS),’’ ’’partially satisfied (PS),’’ ’’not satisfied (NS),’’ and ’’not applicable (NA).’’ The results showing a prevalence of “PS” or “NS” in several items in the Methods section were similar to our findings, although “setting and location” was fully categorized as “FS”, unlike in the present study. In line with the present study, some performed systematic reviews of HEEs examining reporting quality in the fields of cardiology, neurology, plastic surgery, and artificial intelligence in healthcare.^19–22^ Some of them also reported adherence to each item of CHEERS 2013, and similar trends, *e.g.*, insufficient reporting in the Methods section, such as the study by Tai *et al.*, were observed.^20,21^ All of these studies commonly used CHEERS 2013; however, they share common features. First, adherence to each item was dichotomized into binary values of presence or absence. However, as described in the Methods section, some items encompass multiple assessment dimensions, thereby reducing the binary approach to representing the quality of reporting. Second, all studies consistently defined a total score and used CHEERS 2013 as an assessment tool for reporting quality indicators for each included study. However, the CHEERS 2022 statement strongly discourages such an application of CHEERS, noting that CHEERS was not developed as a scoring tool and that such misuse could lead to misleading interpretations of the results.^15^ Our study was carefully designed to address these two concerns commonly observed in previous systematic reviews of reporting quality. In terms of HEEs of ICIs, we found only one systematic review.^23^ However, this systematic review only included HEEs of ICIs published up to April 2018, and only three ICIs were included, *i.e.*, nivolumab, pembrolizumab, and atezolizumab. Moreover, only 30 HEEs were included, representing approximately 12% of our study. The quality of reporting was not assessed, suggesting that the quality of reporting in the HEEs of ICIs remains unclear. Taken together, this is the first study to systematically review HEE studies on ICIs and assess their reporting quality.

Our study has several limitations. First, the results were limited to the HEEs of ICIs and cannot be extrapolated to all anticancer drugs or HEE studies. Second, this study did not examine adherence to or citation of CHEERS 2022. Hence, HEE studies of ICIs published after January 2023 should be assessed for CHEERS 2022 compliance using a methodology similar to that used in this study. Third, this study only assessed the quality of reporting and not the quality of the research methodology itself. An evaluation methodology is essential for policymakers to consider reimbursements and price adjustments. Therefore, future studies may need to assess the methodological quality of HEE studies on ICIs using quality standards such as the CHEQUE tool.^31^

Our study had several strengths. First, a systematic review was conducted using a rigorous methodology, including the development of a prespecified protocol and the registration of PROSPERO. The search strategies were developed by two librarians who used four major databases that were important for a comprehensive search for HEE studies. In addition, screening, data extraction, and analyses were performed according to the methodology of the Cochrane Handbook, which is the current methodological standard for systematic reviews.^32^ Therefore, high reproducibility of the results is expected. Second, the comprehensiveness of the results is expected to be high despite the fact that the search period started in January 2014. This is because the first ICI, nivolumab, was approved by the FDA in December 2014, approximately one year after the start of the search period for this study. This may be supported by the fact that only one HEE study on ICIs was identified between 2014 and 2015. Third, adding a “partially reported” category to the CHEERS 2013 items is also a strength. This allowed for a clearer understanding of the extent to which the researchers adhered to each item in the CHEERS 2013 statement. In addition, not using the CHEERS statement as a scoring tool, a common form of “misuse” emphasized in CHEERS 2022,^15^ can also be considered a strength.

This study had several implications. First, it highlights the importance of comprehensive reporting of items such as methods, which were frequently underreported in this study, for researchers conducting HEE studies on ICIs. Specifically, “setting and location” was largely underreported in this study, although it was crucial to the transferability of the results. Transparent reporting on this point is also essential for policymakers and stakeholders who refer to ICI HEE studies.

## Conclusion

In this systematic review of 252 HEE studies on ICIs from 2014 to 2022, comprehensive reporting was generally limited to the 24 items included in the CHEERS 2013 statement, regardless of the risk of sponsorship bias or citation of CHEERS 2013. The “setting and location” item in the Methods section was particularly underreported. This study highlights the importance of transparent reporting in HEE studies of ICIs.

## Supporting information

PRISMA checklist

## Author contributions

*Concept and design:* Yoshioka, Azuma, Itaya, and Goto

*Acquisition of data:* Yoshioka, Azuma, Funada, and Itaya

*Analysis and interpretation of data:* Yoshioka, Azuma, Funada, Itaya, and Goto

*Drafting the manuscript:* Yoshioka

*Critical revision of paper for important intellectual content:* Azuma, Funada, Itaya, and Goto

*Statistical analysis:* Azuma

*Provision of study materials or patients:* Yoshioka, Azuma, Funada, and Itaya

*Obtaining funding:* Yoshioka and Goto

*Administrative, technical, or logistic support:* Yoshioka, Funada, Itaya, and Goto

*Supervision:* Funada, Itaya, and Goto

## Acknowledgments

We thank Ms. Izumi Endo and Ms. Asako Nishizaki for their assistance with the search strategies. In addition, we acknowledge the National Cancer Center Research Grant (Grant Number: 2022-A-25) for supporting the English language editing fee and the National Institute of Public Health for supporting the article publication fee.

## Data availability statement

All data produced in this review will be available upon reasonable request to the authors.

