## Appendix for "Reporting Quality in Health Economic Evaluation Studies of Immune Checkpoint Inhibitors: A Systematic Review"

**Appendix Table 1 Detailed search strategies for systematic review.**

Database: MEDLINE (PubMed)

Data Parameters: 01/01/2014 to 12/31/2022

Date Searched: 07/07/2023

| **#** | **Searches** | **Results** |
| --- | --- | --- |
| #1 | Neoplasms[MH] OR neoplasm*[TIAB] OR cancer*[TIAB] OR tumor*[TIAB] OR tumour*[TIAB] OR carcinoma*[TIAB] OR adenocarcinoma*[TIAB] OR "Medical Oncology"[MH] OR leukemi*[TIAB] OR leukaemi*[TIAB] OR lymphoma*[TIAB] OR malignan*[TIAB] OR oncolog*[TIAB] OR metasta*[TIAB] | 5,176,775 |
| #2 | "Antibodies, Monoclonal"[MH] OR "Antineoplastic Agents, Immunological"[MH] OR checkpoint[TIAB] OR "check point"[TIAB] OR ICIs[TIAB] OR ctla-4[TIAB] OR pd-1[TIAB] OR pd-l1[TIAB] OR Ipilimumab[TW] OR Yervoy[TW] OR Nivolumab[TW] OR Opdivo[TW] OR Pembrolizumab[TW] OR Keytruda[TW] OR Lambrolizumab[TW] OR Atezolizumab[TW] OR Tecentriq[TW] OR Avelumab[TW] OR Bavencio[TW] OR Durvalumab[TW] OR Imfinzi[TW] OR Cemiplimab[TW] OR Libtayo[TW] OR Dostarlimab[TW] OR Jemperli[TW] OR Relatlimab[TW] OR Tremelimumab[TW] OR Imjudo[TW] OR Ticilimumab[TW] OR Retifanlimab[TW] OR Sintilimab[TW] OR Toripalimab[TW] OR Camrelizumab[TW] OR Carrelizumab[TW] OR Tislelizumab[TW] OR Envafolimab[TW] OR Penpulimab[TW] OR Sugemalimab[TW] OR Zimberelimab[TW] OR Cadonilimab[TW] OR Serplulimab[TW] | 362,455 |
| #3 | #1 AND #2 | 177,228 |
| #4 | cost*[TIAB] OR "Costs and cost analysis"[MeSH:noexp] OR cost benefit analys*[TIAB] OR "Cost-Benefit Analysis"[MH] OR "Health care costs"[MeSH:noexp] | 846,320 |
| #6 | #3 AND #4 | 5,590 |
| #7 | #6 AND (2014/01/01[DP]:2022/12/31[DP] AND English[LA]) | 3,491 |

Database: EMBASE (Elsevier)

Data Parameters: 01/01/2014 to 12/31/2022

Date Searched: 07/07/2023

| **#** | **Searches** | **Results** |
| --- | --- | --- |
| #1 | neoplasm'/exp OR 'oncology'/exp OR neoplasm*:ti,ab,kw OR cancer*:ti,ab,kw OR tumor*:ti,ab,kw OR tumour*:ti,ab,kw OR carcinoma*:ti,ab,kw OR adenocarcinoma*:ti,ab,kw OR leukaemi*:ti,ab,kw OR lymphoma*:ti,ab,kw OR malignan*:ti,ab,kw OR oncolog*:ti,ab,kw OR metasta*:ti,ab,kw | 7,421,553 |
| #2 | monoclonal antibody'/exp OR 'immunological antineoplastic agent'/exp OR checkpoint:ti,ab,kw OR 'check point':ti,ab,kw OR icis:ti,ab,kw OR 'ctla-4':ti,ab,kw OR 'pd-1':ti,ab,kw OR 'pd-l1':ti,ab,kw OR ipilimumab:ti,ab,kw OR yervoy:ti,ab,kw OR nivolumab:ti,ab,kw OR opdivo:ti,ab,kw OR pembrolizumab:ti,ab,kw OR keytruda:ti,ab,kw OR lambrolizumab:ti,ab,kw OR atezolizumab:ti,ab,kw OR tecentriq:ti,ab,kw OR avelumab:ti,ab,kw OR bavencio:ti,ab,kw OR durvalumab:ti,ab,kw OR imfinzi:ti,ab,kw OR cemiplimab:ti,ab,kw OR libtayo:ti,ab,kw OR dostarlimab:ti,ab,kw OR jemperli:ti,ab,kw OR relatlimab:ti,ab,kw OR tremelimumab:ti,ab,kw OR Imjudo:ti,ab,kw OR ticilimumab:ti,ab,kw OR retifanlimab:ti,ab,kw OR sintilimab:ti,ab,kw OR toripalimab:ti,ab,kw OR camrelizumab:ti,ab,kw OR carrelizumab:ti,ab,kw OR tislelizumab:ti,ab,kw OR envafolimab:ti,ab,kw OR penpulimab:ti,ab,kw OR sugemalimab:ti,ab,kw OR zimberelimab:ti,ab,kw OR cadonilimab:ti,ab,kw OR serplulimab:ti,ab,kw | 863,438 |
| #3 | #1 AND #2 | 480,900 |
| #4 | cost'/exp OR 'cost benefit analysis'/exp OR cost*:ti,ab,kw | 1,284,102 |
| #5 | #3 AND #4 | 21,646 |
| #6 | #5 AND ([2014-2022]/py) | 13,435 |
| #7 | #6 AND (English:la) | 13,255 |
| #8 | #7 NOT [conference abstract]/lim | 7,239 |
| #9 | #8 AND [embase]/lim NOT ([embase]/lim AND [medline]/lim) | 1,858 |

Database: CENTRAL (the Cochrane Library)

Data Parameters: 01/01/2014 to 12/31/2022

Date Searched: 07/07/2023

| **#** | **Searches** | **Results** |
| --- | --- | --- |
| #1 | [mh Neoplasms] OR [mh "Medical Oncology"] OR (neoplasm* OR cancer* OR tumor* OR tumour* OR carcinoma* OR adenocarcinoma* OR leukemi* OR leukaemi* OR lymphoma* OR malignan* OR oncolog* OR metasta*):ti,ab,kw | 283,178 |
| #2 | [mh "Antibodies, Monoclonal"] OR [mh "Antineoplastic Agents, Immunological"] OR (checkpoint OR "check point" OR ICIs OR ctla-4 OR pd-1 OR pd-l1 OR Ipilimumab OR Yervoy OR Nivolumab OR Opdivo OR Pembrolizumab OR Keytruda OR Lambrolizumab OR Atezolizumab OR Tecentriq OR Avelumab OR Bavencio OR Durvalumab OR Imfinzi OR Cemiplimab OR Libtayo OR Dostarlimab OR Jemperli OR Relatlimab OR Tremelimumab OR Imjudo OR Ticilimumab OR Retifanlimab OR Sintilimab OR Toripalimab OR Camrelizumab OR Carrelizumab OR Tislelizumab OR Envafolimab OR Penpulimab OR Sugemalimab OR Zimberelimab OR Cadonilimab OR Serplulimab):ti,ab,kw | 27,746 |
| #3 | #1 AND #2 | 18,152 |
| #4 | [mh ^"Costs and cost analysis"] OR [mh "Cost-Benefit Analysis"] OR [mh ^"Health care costs"] OR (cost* OR "cost benefit analysis"):ti,ab,kw | 85,734 |
| #5 | #3 AND #4 | 683 |
| #6 | with Publication Year from 2014 to 2022, in Trials | 551 |
| #7 | #6 NOT Pubmed:an | 340 |

Database: HTA Database (INAHTA)

Data Parameters: 01/01/2014 to 12/31/2022

Date Searched: 07/07/2023

| **#** | **Searches** | **Results** |
| --- | --- | --- |
| #1 | checkpoint OR (check point) OR ICIs OR ctla-4 OR pd-1 OR pd-l1 OR Ipilimumab OR Yervoy OR Nivolumab OR Opdivo OR Pembrolizumab OR Keytruda OR Lambrolizumab OR Atezolizumab OR Tecentriq OR Avelumab OR Bavencio OR Durvalumab OR Imfinzi OR Cemiplimab OR Libtayo OR Dostarlimab OR Jemperli OR Relatlimab OR Tremelimumab OR Imjudo OR Ticilimumab OR Retifanlimab OR Sintilimab OR Toripalimab OR Camrelizumab OR Carrelizumab OR Tislelizumab OR Envafolimab OR Penpulimab OR Sugemalimab OR Zimberelimab OR Cadonilimab OR Serplulimab | 231 |

**Appendix Table 2 Checklist for reporting quality based on the CHEERS 2013 statement.**

**Section/Item/Recommendations**

**Title and abstract**

**1. Title**

a) The study is identified as an economic evaluation study, specifically using terms such as "cost-effectiveness analysis."

b) The evaluated interventions are included.

**2. Abstract**

a) The objectives are stated.

b) The perspective is stated.

c) The setting is stated.

d) The methods (including study design and inputs) are stated.

e) The results (including base-case analysis and uncertainty analysis) are stated.

f) The conclusions are stated.

**Introduction**

**3. Background and objectives**

a) The broad background of the study is clearly stated.

b) The research question and its relationship to healthcare policy or clinical practice are stated.

**Methods**

**4. Target population and subgroups**

a) The characteristics of the base-case population are stated.

b) The reasons for selecting the base-case population are stated.

c) (If applicable) The characteristics of the subgroups in the baseline population are stated.

d) (If applicable) The reasons for selecting the subgroups in the baseline population are stated.

**5. Setting and location**

a) The relevant aspects of the systems in which decision-making must be made are stated.

**6. Study perspective**

a) The perspective of the study is stated.

b) The costs being evaluated are stated in relation to the perspective.

**7. Comparators**

a) The interventions or strategies being compared are stated.

b) The reasons for selecting these comparators are stated.

**8. Time horizon**

a) The time horizons for evaluating costs and outcomes are stated.

b) The reasons why the time horizons for evaluating costs and outcomes are appropriate are stated.

**9. Discount rate**

a) The discount rates for costs and outcomes are stated.

b) The reasons why the values used for the discount rates of costs and outcomes are appropriate are stated.

**10. Choice of health outcomes**

a) The outcomes used as benefit measures in the evaluation are stated.

b) The validity of the type of analysis performed (reasons for selecting the outcomes) is stated.

**11a. Measurement of effectiveness (Single study-based estimates)**

a) The design features of the single effectiveness study are stated.

b) The reasons why the single study was a sufficient source of clinical effectiveness data are stated.

**11b. Measurement of effectiveness (Synthesis-based estimates)**

a) The methods for identifying the studies included for synthesis are stated.

b) The methods for synthesizing clinical effectiveness data are stated.

**12. Measurement and valuation of preference-based outcomes**

a) (If applicable) The population from which outcome preferences were elicited is stated.

b) (If applicable) The methods used for eliciting outcome preferences are stated.

**13a. Estimating resources and costs (Single study-based economic evaluation)**

a) The methods for estimating resource consumption associated with alternative interventions are stated.

b) The methods for valuing each resource consumption item regarding unit cost in primary or secondary research are stated.

c) (If applicable) Any adjustments to approximate opportunity costs are stated.

**13b. Estimating resources and costs (Model-based economic evaluation)**

a) The methods for estimating resource consumption associated with model health states are stated.

b) The data sources for estimating resource consumption associated with model health states are stated.

c) The methods for valuing each resource consumption item regarding unit cost in primary or secondary research are stated.

d) (If applicable) Any adjustments to approximate opportunity costs are stated.

**14. Currency, price date, and conversion**

a) The dates when resource consumptions were estimated are stated.

b) The dates when unit costs were estimated are stated.

c) (If applicable) The methods for adjusting estimated unit costs to the reported cost year are stated.

d) (If applicable) The methods for converting costs into a common currency and the exchange rates are stated.

**15. Choice of model (Model-based economic evaluation)**

a) The specific type of decision-analytic model is stated.

b) The reasons for selecting this specific model type are stated.

c) The structure of the model is shown in a diagram.

**16. Assumptions (Model-based economic evaluation)**

a) All structural or other assumptions necessary for the decision-analytic model are stated.

b) The data sources for each assumption are stated.

**17a. Analytic methods (Single study-based economic evaluation)**

a) The methods and results of regression models used to adjust for differences in costs, outcomes, and cost-effectiveness that can be explained by variability among patient subgroups are stated.

b) The methods for handling uncertainty are stated.

c) The methods for handling data (such as skewed, missing, or censored) are stated.

d) The methods for examining or adjusting the model's validity are stated.

e) (If applicable) The methods for handling population heterogeneity are stated.

**17b. Analytic methods (Model-based economic evaluation)**

a) The methods used to estimate parameters are stated.

b) The methods for handling uncertainty are stated.

c) The methods for handling data (such as skewed, missing, or censored) are stated.

d) The extrapolation methods are stated.

e) The methods for synthesizing data are stated.

f) The methods for examining or adjusting the model's validity are stated.

g) (If applicable) The methods for handling population heterogeneity are stated.

**Results**

**18. Study parameters**

a) The values for all parameters are stated.

b) The ranges for all parameters are stated.

c) The references for all parameters are stated.

d) The input values are presented in tabular form.

e) (If applicable) The probability distributions for all parameters are stated.

f) (If applicable) The rationale and sources for the distributions used to represent uncertainty are stated.

**19. Incremental costs and outcomes**

a) The mean values of estimated interest costs for each intervention are stated.

b) The mean values of estimated outcomes of interest for each intervention are stated.

c) The mean differences (incremental costs) between the comparator groups for each intervention are stated.

d) The mean differences (incremental outcomes) between the comparator groups for each intervention are stated.

e) The incremental cost-effectiveness ratios or whether it is dominant or dominated for each intervention are stated.

**20a. Characterizing uncertainty (Single study-based economic evaluation)**

a) The effects of sampling uncertainty for estimated incremental cost, incremental effectiveness, and incremental cost-effectiveness are stated.

b) The impact of methodological assumptions (such as discount rate and study perspective) for estimated incremental cost, incremental effectiveness, and incremental cost-effectiveness are stated.

**20b. Characterizing uncertainty (Model-based economic evaluation)**

a) The effects on the uncertainty result for all input parameters are stated.

b) The effects on the results of uncertainty related to the structure of the model are stated.

c) The effects on the results of uncertainty related to the assumptions of the model are stated.

**21. Characterizing heterogeneity**

a) (If applicable) The differences in costs that can be explained by variability between subgroups of patients with different baseline characteristics are stated.

b) (If applicable) The differences in outcomes that can be explained by variability between subgroups of patients with different baseline characteristics are stated.

c) (If applicable) The differences in cost-effectiveness that can be explained by variability between subgroups of patients with different baseline characteristics are stated.

d) (If applicable) Other observed variability in effects that are not reducible by more information are stated.

**Discussion**

**22. Study findings, limitations, generalizability, and current knowledge**

a) Key study findings are summarized, and how they support the conclusions is stated.

b) The limitations of the findings are stated.

c) The generalizability of the findings is stated.

d) How the findings fit with current knowledge is stated.

**Other**

**23. Source of funding**

a) The source of funding for the study is stated.

b) Other non-monetary sources of support are stated.

c) The role of the funders (identification, design, conduct, and reporting of the analysis) is stated.

**24. Conflicts of interest**

a) The presence or absence of conflicts of interest is stated.

**Appendix Table 3 Summary of the included health economic evaluation studies of immune checkpoint inhibitors.**

| Author and Year | Setting | Study design | Target population | Intervention | Study perspective | Time horizon | Risk of sponsorship bias | Citation of the CHEERS 2013 statement |
| --- | --- | --- | --- | --- | --- | --- | --- | --- |
| Curl et al., 2014^1^ | US | CUA | Melanoma | Ipilimumab + Vemurafenib | Societal perspective | Lifetime | Absent | Not cited |
| Bohensky et al., 2016^2^ | Australia | CEA/CUA | Melanoma | Nivolumab | Health system or payer perspective | 10 years | Present | Not cited |
| De Francesco et al., 2016^3^ | Italy | CEA/CUA | Melanoma | Ipilimumab | Health system or payer perspective | 15 years | Present | Not cited |
| Jensen, I.S., 2016^4^ | US | CBA | Melanoma | Ipilimumab + Nivolumab | Payer perspective; Societal perspective | Not reported | Present | Not cited |
| Huang et al., 2017^5^ | US | CEA/CUA | Non-small cell lung cancer | Pembrolizumab | Health system or payer perspective | 20 years | Present | Not cited |
| Huang et al., 2017^6^ | US | CEA/CUA | Non-small cell lung cancer | Pembrolizumab | Health system or payer perspective | Lifetime (20 years) | Present | Not cited |
| Kohn et al., 2017^7^ | US | CUA | Melanoma | Nivolumab | Health system or payer perspective | Lifetime | Absent | Not cited |
| Miguel et al., 2017^8^ | Portugal | CEA/CUA | Melanoma | Pembrolizumab | Health system or payer perspective | Lifetime (40 years) | Present | Not cited |
| Oh et al., 2017^9^ | US | CUA | Melanoma | Ipilimumab + Nivolumab | Societal perspective | 175 months | Absent | Cited |
| Pike et al., 2017^10^ | Norway | CUA | Melanoma | Nivolumab; Pembrolizumab | Health system or payer perspective | 10 years | Absent | Not cited |
| Wan et al., 2017^11^ | China and US | CEA/CUA | Renal cell carcinoma | Nivolumab | Health system or payer perspective | 20 years | Absent | Not cited |
| Wang et al., 2017^12^ | US | CEA/CUA | Melanoma | Pembrolizumab | Health system or payer perspective | 20 years | Present | Cited |
| Ward et al., 2017^13^ | US | CUA | Head and neck cancer | Nivolumab | Health system or payer perspective | 36 months | Absent | Cited |
| Aguiar Jr et al., 2018^14^ | Argentina, Brazil, and Peru | CEA/CUA | Non-small cell lung cancer | Pembrolizumab | Health system or payer perspective | 5 years | Present | Not cited |
| Georgieva et al., 2018^15^ | UK and US | CUA | Non-small cell lung cancer | Pembrolizumab | Health system or payer perspective | Lifetime | Present | Not cited |
| Hu et al., 2018^16^ | UK | CEA/CUA | Non-small cell lung cancer | Pembrolizumab | Health system or payer perspective | Not reported | Absent | Not cited |
| Insinga et al., 2018^17^ | US | CEA/CUA | Non-small cell lung cancer | Pembrolizumab + Chemotherapy | Health system or payer perspective | 20 years | Present | Not cited |
| Large et al., 2018^18^ | US | CEA/CUA | Hodgkin lymphoma | Pembrolizumab | Health system or payer perspective | 20 years | Present | Not cited |
| McCrea et al., 2018^19^ | US | CEA/CUA | Renal cell carcinoma | Nivolumab | Health system or payer perspective | Lifetime (25 years) | Present | Not cited |
| Meng et al., 2018^20^ | UK | CEA/CUA | Renal cell carcinoma | Nivolumab | Health system or payer perspective | 30 years | Present | Not cited |
| Meng et al., 2018^21^ | UK | CEA/CUA | Melanoma | Nivolumab | Not reported | Not reported | Present | Not cited |
| Raphael et al., 2018^22^ | Canada | CBA/CEA/CUA | Renal cell carcinoma | Nivolumab | Health system or payer perspective | Lifetime | Absent | Not cited |
| Sarfaty et al., 2018^23^ | Australia, Canada, UK, and US | CUA | Urothelial carcinoma (Bladder cancer) | Pembrolizumab | Health system or payer perspective | 5 years | Absent | Not cited |
| Sarfaty et al., 2018^24^ | US | CEA/CUA | Renal cell carcinoma | Nivolumab | Health system or payer perspective | 10 years | Absent | Not cited |
| Tringale et al., 2018^25^ | Not reported | CUA | Head and neck cancer | Nivolumab | Health system or payer perspective | 30 years | Absent | Not cited |
| Wu et al., 2018^26^ | China, UK, and US | CEA/CUA | Renal cell carcinoma | Ipilimumab + Nivolumab | Health system or payer perspective | 10 years | Absent | Not cited |
| Zargar et al., 2018^27^ | Canada | CUA | Head and neck cancer | Nivolumab | Health system or payer perspective | 5 years | Absent | Cited |
| Almutairi et al., 2019^28^ | US | CEA/CUA | Melanoma | Ipilimumab + Talimogene Laherparepvec | Health system or payer perspective | Lifetime | Present | Not cited |
| Barrington et al., 2019^29^ | US | CEA | Endometrial carcinoma | Pembrolizumab | Not reported | Not reported | Absent | Not cited |
| Bensimon et al., 2019^30^ | US | CEA/CUA | Melanoma | Pembrolizumab | Health system or payer perspective | Lifetime (46 years) | Present | Not cited |
| Bhadhuri et al., 2019^31^ | Switzerland | CEA/CUA | Non-small cell lung cancer | Pembrolizumab | Health system or payer perspective | 20 years | Present | Not cited |
| Bullement et al., 2019^32^ | UK | CEA/CUA | Merkel cell carcinoma | Avelumab | Health system or payer perspective | Lifetime (40 years) | Present | Not cited |
| Chen et al., 2019^33^ | China | CEA/CUA | Renal cell carcinoma | Pembrolizumab + Axitinib | Health system or payer perspective | Lifetime | Absent | Not cited |
| Chouaid et al., 2019^34^ | France | CEA/CUA | Non-small cell lung cancer | Pembrolizumab | Health system or payer perspective | 10 years | Present | Not cited |
| Chu et al., 2019^35^ | US | CEA/CUA | Colorectal cancer | Ipilimumab + Nivolumab; Nivolumab | Health system or payer perspective | Lifetime | Present | Not cited |
| Criss et al., 2019^36^ | US | CEA/CUA | Non-small cell lung cancer | Durvalumab + Chemotherapy | Societal perspective | 5 years | Present | Cited |
| Criss et al., 2019^37^ | US | CUA | Non-small cell lung cancer | Atezolizumab + Bevacizumab + Carboplatin + Paclitaxel | Health care sector perspective | Not reported | Present | Cited |
| Deniz et al., 2019^38^ | US | CEA | Renal cell carcinoma | Nivolumab | Health system or payer perspective | Lifetime (25 years) | Present | Not cited |
| Gao et al., 2019^39^ | Australia | CEA/CUA | Non-small cell lung cancer | Nivolumab | Health system or payer perspective | 6 years | Absent | Not cited |
| Huang et al., 2019^40^ | US | CEA/CUA | Non-small cell lung cancer | Pembrolizumab | Health system or payer perspective | 20 years | Present | Not cited |
| Insinga et al., 2019^41^ | US | CEA/CUA | Non-small cell lung cancer | Pembrolizumab + Chemotherapy | Health system or payer perspective | 20 years | Present | Not cited |
| Li et al., 2019^42^ | China | CUA | Small cell lung cancer | Atezolizumab + Chemotherapy | Not reported | Not reported | Absent | Not cited |
| Liao et al., 2019^43^ | China | CUA | Non-small cell lung cancer | Pembrolizumab | Health system or payer perspective | 10 years | Absent | Not cited |
| Liu et al., 2019^44^ | China and US | CUA | Head and neck cancer | Pembrolizumab | Health system or payer perspective | 10 years | Absent | Not cited |
| Ondhia et al., 2019^45^ | Canada | CEA/CUA | Non-small cell lung cancer | Atezolizumab; Nivolumab | Health system or payer perspective | 10 years | Present | Not cited |
| Patterson et al., 2019^46^ | Sweden | CEA/CUA | Urothelial carcinoma | Pembrolizumab | Health system or payer perspective | Lifetime (15 years) | Present | Not cited |
| Quon et al., 2019^47^ | Canada | CEA/CUA | Melanoma | Ipilimumab + Nivolumab | Health system or payer perspective | 20 years | Present | Not cited |
| Reinhorn et al., 2019^48^ | US | CUA | Renal cell carcinoma | Ipilimumab + Nivolumab | Health system or payer perspective | 10 years | Absent | Not cited |
| She et al., 2019^49^ | US | CEA/CUA | Non-small cell lung cancer | Pembrolizumab | Health system or payer perspective | 20 years | Absent | Not cited |
| Wan et al., 2019^50^ | US | CEA/CUA | Non-small cell lung cancer | Atezolizumab + Bevacizumab + Carboplatin + Paclitaxel | Health system or payer perspective | Lifetime | Absent | Not cited |
| Wan et al., 2019^51^ | US | CEA/CUA | Renal cell carcinoma | Ipilimumab + Nivolumab | Health system or payer perspective | Lifetime | Absent | Not cited |
| Zeng et al., 2019^52^ | US | CUA | Non-small cell lung cancer | Pembrolizumab + Chemotherapy | Health system or payer perspective | 20 years | Absent | Not cited |
| Zhou et al., 2019^53^ | China | CUA | Non-small cell lung cancer | Pembrolizumab | Health system or payer perspective | 10 years | Absent | Not cited |
| Zhou et al., 2019^54^ | US | CUA | Small cell lung cancer | Atezolizumab + Chemotherapy | American perspective | Not reported | Absent | Not cited |
| Ambavane et al., 2020^55^ | US | CEA/CUA | Renal cell carcinoma | Ipilimumab + Nivolumab | Health system or payer perspective | Lifetime (40 years) | Present | Not cited |
| Armeni et al., 2020^56^ | Italy | CBA/CEA/CUA | Non-small cell lung cancer | Durvalumab | Health system or payer perspective | Lifetime (40 years) | Present | Not cited |
| Aziz et al., 2020^57^ | Singapore | CEA/CUA | Non-small cell lung cancer | Pembrolizumab | Health system or payer perspective | 10 years | Absent | Not cited |
| Bensimon et al., 2020^58^ | US | CEA/CUA | Melanoma | Pembrolizumab | Health system or payer perspective | Lifetime | Present | Not cited |
| Bensimon et al., 2020^59^ | US | CEA/CUA | Renal cell carcinoma | Pembrolizumab + Axitinib | Health system or payer perspective | Lifetime | Present | Not cited |
| Bregman et al., 2020^60^ | France | CEA/CUA | Melanoma | Pembrolizumab | Societal perspective | 20 years | Present | Not cited |
| Criss et al., 2020^61^ | Not reported | CUA | Non-small cell lung cancer | Pembrolizumab + Chemotherapy | Health care sector perspective | Not reported | Present | Not cited |
| Ding et al., 2020^62^ | US | CEA/CUA | Non-small cell lung cancer | Atezolizumab + Chemotherapy | Health system or payer perspective | 15 years | Absent | Not cited |
| Gibson et al., 2020^63^ | UK | CEA/CUA | Melanoma | Ipilimumab + Nivolumab | Health system or payer perspective | Lifetime (40 years) | Present | Not cited |
| Haddad et al., 2020^64^ | US | CEA/CUA | Head and neck cancer | Nivolumab | Health system or payer perspective | 25 years | Present | Not cited |
| Han et al., 2020^65^ | US | CEA/CUA | Non-small cell lung cancer | Durvalumab | Health system or payer perspective | Lifetime | Absent | Not cited |
| Hu et al., 2020^66^ | Not reported | CEA | Melanoma | Pembrolizumab | Not reported | 10 years | Present | Not cited |
| Hu et al., 2020^67^ | US | CEA/CUA | Non-small cell lung cancer | Ipilimumab + Nivolumab | Health system or payer perspective | 20 years | Absent | Not cited |
| Lang et al., 2020^68^ | China and US | CEA/CUA | Head and neck cancer | Pembrolizumab + Chemotherapy | Health system or payer perspective | 20 years | Absent | Not cited |
| Li et al., 2020^69^ | US | CEA/CUA | Non-small cell lung cancer | Ipilimumab + Nivolumab | Health system or payer perspective | 10 years | Absent | Not cited |
| Li et al., 2020^70^ | US | CEA/CUA | Breast cancer | Atezolizumab + nab-Paclitaxel | Health system or payer perspective | 10 years | Absent | Not cited |
| Lin et al., 2020^71^ | US | CEA/CUA | Non-small cell lung cancer | Atezolizumab + Carboplatin + nab-Paclitaxel | Health system or payer perspective | 10 years | Absent | Not cited |
| Liu et al., 2020^72^ | China | CEA/CUA | Non-small cell lung cancer | Nivolumab | Health system or payer perspective | Lifetime | Absent | Not cited |
| Loong et al., 2020^73^ | Hong Kong | CEA/CUA | Melanoma | Pembrolizumab | Health system or payer perspective | 30 years | Present | Not cited |
| Lu et al., 2020^74^ | US | CUA | Renal cell carcinoma | Avelumab + Axitinib | Health system or payer perspective | 10 years | Absent | Cited |
| Marine et al., 2020^75^ | France | CEA/CMA/CUA | Non-small cell lung cancer | Atezolizumab | Health system or payer perspective | 10 years | Present | Not cited |
| Paly et al., 2020^76^ | Japan | CEA/CUA | Melanoma | Ipilimumab + Nivolumab | Health system or payer perspective | 30 years | Present | Not cited |
| Panje et al., 2020^77^ | Switzerland | CEA/CUA | Non-small cell lung cancer | Durvalumab | Health system or payer perspective | 10 years | Present | Cited |
| Parmar et al., 2020^78^ | Canada | CEA/CUA | Urothelial carcinoma (Bladder cancer) | Atezolizumab | Health system or payer perspective | 5 years | Present | Not cited |
| Phua et al., 2020^79^ | Singapore | CEA/CUA | Breast cancer | Atezolizumab + nab-Paclitaxel | Health system or payer perspective | 5 years | Absent | Not cited |
| Sherrow et al., 2020^80^ | US | CUA | Hepatocellular carcinoma | Pembrolizumab | Not reported | Not reported | Absent | Not cited |
| Slater et al., 2020^81^ | US | CEA/CUA | Urothelial carcinoma | Pembrolizumab | Health system or payer perspective | 20 years | Present | Not cited |
| Srivastava et al., 2020^82^ | Sweden | CEA/CUA | Urothelial carcinoma | Pembrolizumab | Health system or payer perspective | 15 years | Present | Not cited |
| Wan et al., 2020^83^ | China and US | CUA | Non-small cell lung cancer | Pembrolizumab + Chemotherapy | Health system or payer perspective | Lifetime | Absent | Not cited |
| Watson et al., 2020^84^ | US | CUA | Renal cell carcinoma | Pembrolizumab + Axitinib | Health care sector perspective | Not reported | Absent | Cited |
| Weng et al., 2020^85^ | US | CUA | Non-small cell lung cancer | Pembrolizumab | Health system or payer perspective | Lifetime | Absent | Not cited |
| Weng et al., 2020^86^ | China and US | CEA/CUA | Breast cancer | Atezolizumab + nab-Paclitaxel | Health system or payer perspective | Lifetime | Absent | Not cited |
| Wu et al., 2020^87^ | US | CBA/CEA/CUA | Breast cancer | Atezolizumab + nab-Paclitaxel | Health system or payer perspective | 10 years | Absent | Not cited |
| Wu et al., 2020^88^ | US | CBA/CEA/CUA | Melanoma | Ipilimumab + Nivolumab; Pembrolizumab | Health system or payer perspective | Lifetime | Absent | Not cited |
| Xin et al., 2020^89^ | China | CUA | Head and neck cancer | Pembrolizumab | Health system or payer perspective | 30 years | Absent | Not cited |
| Yeh et al., 2020^90^ | Not reported | CUA | Head and neck cancer | Nivolumab; Pembrolizumab | Health system or payer perspective | Not reported | Absent | Not cited |
| Zhang et al., 2020^91^ | China | CEA/CUA | Non-small cell lung cancer | Nivolumab | Health system or payer perspective | 10 years | Absent | Not cited |
| Zhang et al., 2020^92^ | US | CEA/CUA | Small cell lung cancer | Durvalumab + Etoposide + Platinum | Health system or payer perspective | 10 years | Absent | Not cited |
| Zhang et al., 2020^93^ | China | CUA | Esophageal cancer | Nivolumab | Societal perspective | 10 years | Absent | Not cited |
| Zhou et al., 2020^94^ | China | CUA | Head and neck cancer | Pembrolizumab | Health system or payer perspective | 10 years | Absent | Not cited |
| Zhu et al., 2020^95^ | US | CUA | Renal cell carcinoma | Pembrolizumab + Axitinib | Health system or payer perspective | 20 years | Absent | Not cited |
| Ackroyd et al., 2021^96^ | US | CUA | Endometrial carcinoma | Pembrolizumab + Lenvatinib | Health system or payer perspective | 36 months | Absent | Not cited |
| Barbier et al., 2021^97^ | Switzerland | CEA/CUA | Non-small cell lung cancer | Pembrolizumab | Health system or payer perspective | 10 years | Absent | Not cited |
| Barrington et al., 2021^98^ | US | CUA | Endometrial carcinoma | Pembrolizumab + Lenvatinib | Not reported | Not reported | Absent | Not cited |
| Cai et al., 2021^99^ | US | CEA/CUA | Melanoma | Atezolizumab + Cobimetinib + Vemurafenib | Health system or payer perspective | Lifetime (30 years) | Present | Cited |
| Cai et al., 2021^100^ | China | CEA/CUA | Esophageal cancer | Camrelizumab | Not reported | 10 years | Absent | Not cited |
| Cai et al., 2021^101^ | China | CUA | Non-small cell lung cancer | Pembrolizumab + Chemotherapy | Health system or payer perspective | 10 years | Absent | Not cited |
| Chang et al., 2021^102^ | Taiwan | CEA/CUA | Merkel cell carcinoma | Avelumab | Not reported | Lifetime (40 years) | Present | Cited |
| Chaudhary et al., 2021^103^ | US | CEA/CUA | Non-small cell lung cancer | Nivolumab | Health system or payer perspective | Lifetime | Present | Not cited |
| Chaudhary et al., 2021^104^ | Canada and Sweden | CEA/CUA | Non-small cell lung cancer | Nivolumab | Health system or payer perspective | 10 years (CAN); 15 years (SWE) | Present | Not cited |
| Cheng et al., 2021^105^ | China and US | CEA/CUA | Non-small cell lung cancer | Atezolizumab | Health system or payer perspective | 20 years | Absent | Cited |
| Chiang et al., 2021^106^ | US | CEA/CUA | Hepatocellular carcinoma | Pembrolizumab | Health system or payer perspective | 3 years | Present | Cited |
| Chiang et al., 2021^107^ | US | CEA/CUA | Hepatocellular carcinoma | Atezolizumab + Bevacizumab | Health system or payer perspective | 5 years | Absent | Cited |
| Chisaki et al., 2021^108^ | Japan | CEA/CUA | Breast cancer | Atezolizumab + nab-Paclitaxel | Health system or payer perspective | 3 years | Absent | Not cited |
| Chongqing et al., 2021^109^ | US | CEA/CUA | Colorectal cancer | Pembrolizumab | Health system or payer perspective | Lifetime | Absent | Not cited |
| Courtney et al., 2021^110^ | US | CUA | Non-small cell lung cancer | Ipilimumab + Nivolumab | Health system or payer perspective | 10 years | Absent | Cited |
| Ding et al., 2021^111^ | US | CEA/CUA | Renal cell carcinoma | Pembrolizumab + Axitinib | Health system or payer perspective | Lifetime | Absent | Not cited |
| Ding et al., 2021^112^ | US | CEA/CUA | Small cell lung cancer | Durvalumab + Chemotherapy | Health system or payer perspective | Lifetime | Absent | Not cited |
| Ghetti et al., 2021^113^ | Italy | CEA/CUA | Cutaneous squamous cell carcinoma | Cemiplimab | Health system or payer perspective | Lifetime (30 years) | Present | Not cited |
| Gil-Rojas et al., 2021^114^ | Colombia | CBA/CEA/CUA | Melanoma | Ipilimumab + Nivolumab | Health system or payer perspective | 22 years | Present | Not cited |
| Hale et al., 2021^115^ | US | CEA/CUA | Urothelial carcinoma | Pembrolizumab | Health system or payer perspective | Lifetime (20 years) | Present | Not cited |
| Hao et al., 2021^116^ | China and US | CBA/CEA/CUA | Non-small cell lung cancer | Ipilimumab + Nivolumab | Health system or payer perspective | 10 years | Absent | Not cited |
| Insinga et al., 2021^117^ | US | CEA/CUA | Non-small cell lung cancer | Pembrolizumab + Chemotherapy | Health system or payer perspective | 20 years | Present | Not cited |
| Kang et al., 2021^118^ | China | CBA/CEA/CUA | Small cell lung cancer | Atezolizumab + Chemotherapy; Durvalumab + Chemotherapy; Ipilimumab + Chemotherapy; Nivolumab + Chemotherapy | Health system or payer perspective | 10 years | Absent | Not cited |
| Khaki et al., 2021^119^ | Not reported | CEA | Urothelial carcinoma (Bladder cancer) | Atezolizumab; Ipilimumab + Nivolumab; Pembrolizumab | Health system or payer perspective | 2 years | Present | Cited |
| Kim et al., 2021^120^ | Republic of Korea | CEA/CUA | Renal cell carcinoma | Nivolumab | Societal perspective | 30 years | Present | Not cited |
| Kim et al., 2021^121^ | Australia | CEA/CUA | Renal cell carcinoma | Nivolumab | Health system or payer perspective | 60 months; 110 months | Present | Not cited |
| Konidaris et al., 2021^122^ | US | CEA/CUA | Cutaneous squamous cell carcinoma | Cemiplimab | Health system or payer perspective | Lifetime (30 years) | Present | Not cited |
| Li et al., 2021^123^ | US | CEA/CUA | Renal cell carcinoma | Nivolumab + Cabozantinib | Health system or payer perspective | Lifetime | Absent | Not cited |
| Li et al., 2021^124^ | US | CEA/CUA | Renal cell carcinoma | Nivolumab + Cabozantinib; Pembrolizumab + Axitinib | Health system or payer perspective | Lifetime | Absent | Not cited |
| Liao et al., 2021^125^ | US | CUA | Renal cell carcinoma | Nivolumab + Cabozantinib | Health system or payer perspective | Lifetime | Absent | Not cited |
| Lin et al., 2021^126^ | China | CEA/CUA | Esophageal cancer | Camrelizumab | Health system or payer perspective | 2 years | Absent | Not cited |
| Lin et al., 2021^127^ | US | CEA/CUA | Small cell lung cancer | Durvalumab + Carboplatin/Cisplatin + Etoposide | Health system or payer perspective | Lifetime | Absent | Not cited |
| Liu et al., 2021^128^ | US | CUA | Small cell lung cancer | Durvalumab + Etoposide + Platinum | Health system or payer perspective | 10 years | Absent | Cited |
| Liu et al., 2021^129^ | China | CEA/CUA | Non-small cell lung cancer | Atezolizumab | Health care sector perspective | 10 years | Absent | Not cited |
| Liu et al., 2021^130^ | US | CEA/CUA | Small cell lung cancer | Pembrolizumab + Etoposide + Platinum | Health system or payer perspective | 10 years | Absent | Not cited |
| Liu et al., 2021^131^ | US | CUA | Non-small cell lung cancer | Pembrolizumab + Chemotherapy | Health system or payer perspective | 20 years | Absent | Cited |
| Liu et al., 2021^132^ | US | CUA | Non-small cell lung cancer | Cemiplimab | Health care sector perspective | Lifetime | Absent | Cited |
| Liu et al., 2021^133^ | China | CEA/CUA | Breast cancer | Atezolizumab + nab-Paclitaxel | Health care sector perspective | 10 years | Absent | Not cited |
| Mehra et al., 2021^134^ | US | CEA/CUA | Non-small cell lung cancer | Durvalumab | Medicare perspective; Commercial insurance perspective | Lifetime (30 years) | Present | Not cited |
| Mojtahed et al., 2021^135^ | US | CEA/CUA | Melanoma | Pembrolizumab | Health system or payer perspective | Lifetime (19.4 years) | Absent | Cited |
| Mulder et al., 2021^136^ | the Netherlands | CEA/CUA | Melanoma | Nivolumab; Pembrolizumab | Societal perspective | Lifetime | Present | Not cited |
| Paul et al., 2021^137^ | US | CEA/CUA | Cutaneous squamous cell carcinoma | Cemiplimab | US perspective | Lifetime (30 years) | Present | Not cited |
| Pei et al., 2021^138^ | US | CEA/CUA | Head and neck cancer | Nivolumab | Health system or payer perspective | 15 years | Absent | Cited |
| Peng et al., 2021^139^ | US | CEA/CUA | Melanoma | Ipilimumab + Nivolumab/Pembrolizumab | Health system or payer perspective | Lifetime | Absent | Not cited |
| Peng et al., 2021^140^ | US | CBA/CEA/CUA | Urothelial carcinoma | Avelumab | Health system or payer perspective | Lifetime | Absent | Not cited |
| Peng et al., 2021^141^ | US | CBA/CEA/CUA | Non-small cell lung cancer | Ipilimumab + Nivolumab + Chemotherapy | Health system or payer perspective | Lifetime | Absent | Not cited |
| Peng et al., 2021^142^ | US | CEA/CUA | Non-small cell lung cancer | Atezolizumab | Health system or payer perspective | Lifetime | Absent | Not cited |
| Qiao et al., 2021^143^ | US | CMA | Non-small cell lung cancer | Pembrolizumab | Health system or payer perspective | 1 year; 2 years; 3 years | Present | Not cited |
| Qiao et al., 2021^144^ | China | CUA | Non-small cell lung cancer | Pembrolizumab + Platinum | Health system or payer perspective | 30 years | Absent | Not cited |
| Qin et al., 2021^145^ | US | CEA/CUA | Urothelial carcinoma | Atezolizumab + Carboplatin/Cisplatin + Gemcitabine | Health system or payer perspective | Lifetime | Absent | Not cited |
| Roth et al., 2021^146^ | US | CEA/CUA | Small cell lung cancer | Nivolumab | Health system or payer perspective | Lifetime (20 years) | Present | Not cited |
| Rothwell et al., 2021^147^ | UK | CEA/CUA | Non-small cell lung cancer | Nivolumab | Health system or payer perspective | 20 years | Present | Not cited |
| Salans et al., 2021^148^ | Not reported | CUA | Melanoma | Ipilimumab | Health system or payer perspective | 10 years | Absent | Not cited |
| Shay et al., 2021^149^ | US | CBA/CUA | Renal cell carcinoma | Ipilimumab + Nivolumab | Health system or payer perspective | 10 years | Present | Not cited |
| Shi et al., 2021^150^ | China | CEA/CUA | Non-small cell lung cancer | Pembrolizumab | Health system or payer perspective | Lifetime | Absent | Cited |
| Smare et al., 2021^151^ | US | CEA/CUA | Small cell lung cancer | Nivolumab | Health system or payer perspective | Lifetime (20 years) | Present | Not cited |
| Standage et al., 2021^152^ | Not reported | CUA | Melanoma | Pembrolizumab | Societal perspective | 5 years | Present | Not cited |
| Su et al., 2021^153^ | US | CBA/CEA/CUA | Hepatocellular carcinoma | Atezolizumab + Bevacizumab | Health system or payer perspective | 2 years | Absent | Cited |
| Takushima et al., 2021^154^ | Japan | CEA/CUA | Gastric cancer | Nivolumab | Health system or payer perspective | 10 years | Present | Not cited |
| Teng et al., 2021^155^ | China | CUA | Non-small cell lung cancer | Atezolizumab; Durvalumab; Pembrolizumab | Health system or payer perspective | Lifetime | Absent | Not cited |
| Thurgar et al., 2021^156^ | US | CEA/CUA | Endometrial carcinoma | Pembrolizumab | Health system or payer perspective | Lifetime (30 years) | Present | Not cited |
| Wan et al., 2021^157^ | US | CEA/CUA | Non-small cell lung cancer | Ipilimumab + Nivolumab | Health system or payer perspective | Lifetime | Absent | Not cited |
| Wang et al., 2021^158^ | US | CEA/CUA | Non-small cell lung cancer | Cemiplimab | Health system or payer perspective | 20 years | Absent | Not cited |
| Wang et al., 2021^159^ | US | CEA/CUA | Small cell lung cancer | Atezolizumab + Chemotherapy | Health system or payer perspective | Lifetime (2.5 years) | Absent | Not cited |
| Wen et al., 2021^160^ | China and US | CEA/CUA | Hepatocellular carcinoma | Atezolizumab + Bevacizumab | Health system or payer perspective | 10 years | Absent | Not cited |
| Wurcel et al., 2021^161^ | Argentina | CEA/CUA | Melanoma | Pembrolizumab | Health system or payer perspective | Lifetime (46 years) | Present | Not cited |
| Wurcel et al., 2021^162^ | Argentina | CEA/CUA | Head and neck cancer | Pembrolizumab + 5-Fluorouracil + Platinum | Societal perspective | Lifetime (20 years) | Present | Not cited |
| Wymer et al., 2021^163^ | US | CUA | Urothelial carcinoma (Bacillus Calmette-Guerin-unresponsive carcinoma in situ) | Pembrolizumab | Health system or payer perspective | 5 years | Present | Not cited |
| Xiang et al., 2021^164^ | China | CEA/CUA | Non-small cell lung cancer | Camrelizumab | Health system or payer perspective | Lifetime | Absent | Not cited |
| Yang et al., 2021^165^ | China | CEA/CUA | Non-small cell lung cancer | Atezolizumab + Chemotherapy | Health system or payer perspective | Lifetime | Absent | Not cited |
| Yang et al., 2021^166^ | US | CEA/CUA | Non-small cell lung cancer | Ipilimumab + Nivolumab; Ipilimumab + Nivolumab + Chemotherapy | Health care sector perspective | Lifetime | Absent | Not cited |
| Yang et al., 2021^167^ | China | CEA/CUA | Esophageal cancer | Camrelizumab | Health system or payer perspective | Lifetime (5 years) | Absent | Cited |
| Zhang et al., 2021^168^ | China | CUA | Esophageal cancer | Camrelizumab + Chemotherapy | Health system or payer perspective | 5 years | Absent | Not cited |
| Zhang et al., 2021^169^ | US | CEA/CUA | Hepatocellular carcinoma | Atezolizumab + Bevacizumab | Health system or payer perspective | 6 years | Absent | Cited |
| Zhu et al., 2021^170^ | China | CEA/CUA | Non-small cell lung cancer | Camrelizumab + Chemotherapy | Health system or payer perspective | Lifetime | Absent | Not cited |
| Zhu et al., 2021^171^ | US | CEA/CUA | Small cell lung cancer | Pembrolizumab + Etoposide + Platinum | Health system or payer perspective | 10 years | Absent | Not cited |
| Aguiar-Ibáñez et al., 2022^172^ | US | CEA/CUA | Colorectal cancer | Pembrolizumab | Health system or payer perspective | Lifetime (40 years) | Present | Not cited |
| Baker et al., 2022^173^ | US | CEA/CUA | Melanoma | Ipilimumab + Nivolumab | Health system or payer perspective | Lifetime (30 years) | Present | Not cited |
| Barrington et al., 2022^174^ | US | CUA | Cervical cancer | Pembrolizumab + Chemotherapy; Pembrolizumab + Bevacizumab + Chemotherapy | Health system or payer perspective | Not reported | Absent | Not cited |
| Berling et al., 2022^175^ | US | CEA/CUA | Non-small cell lung cancer | Ipilimumab + Nivolumab | Health system or payer perspective | Lifetime (20 years) | Present | Not cited |
| Borse et al., 2022^176^ | US | CEA/CUA | Head and neck cancer | Pembrolizumab + 5-Fluorouracil + Platinum | Health system or payer perspective | Lifetime (20 years) | Present | Not cited |
| Buja et al., 2022^177^ | Italy | CEA | Non-small cell lung cancer | Durvalumab | Health system or payer perspective | 5 years | Absent | Not cited |
| Chan et al., 2022^178^ | US | CEA/CUA | Renal cell carcinoma | Pembrolizumab + Axitinib | Health system or payer perspective | Lifetime (20 years) | Absent | Not cited |
| Chen et al., 2022^179^ | China | CEA/CUA | Non-small cell lung cancer | Sugemalimab + Chemotherapy | Health system or payer perspective | 10 years | Absent | Not cited |
| Chen et al., 2022^180^ | China | CEA/CUA | Non-small cell lung cancer | Durvalumab + Chemotherapy | Health system or payer perspective | 10 years | Absent | Not cited |
| Chen et al., 2022^181^ | China | CEA/CUA | Non-small cell lung cancer | Sintilimab + Chemotherapy | Health system or payer perspective | Lifetime | Present | Not cited |
| Chen et al., 2022^182^ | China | CEA/CUA | Non-small cell lung cancer | Atezolizumab | Health system or payer perspective | 10 years | Absent | Not cited |
| Chen et al., 2022^183^ | China | CUA | Non-small cell lung cancer | Camrelizumab + Chemotherapy | Health system or payer perspective | 10 years | Absent | Not cited |
| Feng et al., 2022^184^ | US | CUA | Endometrial carcinoma | Pembrolizumab + Lenvatinib | Health system or payer perspective | Lifetime | Absent | Cited |
| Gong et al., 2022^185^ | China | CEA/CUA | Non-small cell lung cancer | Tislelizumab | Health system or payer perspective | Lifetime (20 years) | Absent | Not cited |
| Hu et al., 2022^186^ | US | CEA/CUA | Esophageal cancer | Pembrolizumab | Health system or payer perspective | 10 years | Absent | Not cited |
| Huang et al., 2022^187^ | US | CEA/CUA | Breast cancer | Pembrolizumab + Chemotherapy | Health system or payer perspective | 20 years | Present | Not cited |
| Ionova et al., 2022^188^ | US | CUA | Small cell lung cancer | Durvalumab | Health system or payer perspective | 360 months | Absent | Not cited |
| Jiang et al., 2022^189^ | China | CEA/CUA | Non-small cell lung cancer | Durvalumab | Health system or payer perspective | Lifetime (40 years) | Absent | Not cited |
| Jiang et al., 2022^190^ | China | CEA/CUA | Non-small cell lung cancer | Pembrolizumab + Carboplatin/Cisplatin + Pemetrexed | Societal perspective | 20 years | Absent | Not cited |
| Jiang et al., 2022^191^ | China | CUA | Gastric cancer, Gastroesophageal junction cancer, and Esophageal adenocarcinoma | Nivolumab + Chemotherapy | Health system or payer perspective | 10 years | Absent | Not cited |
| Kashiwa et al., 2022^192^ | Japan | CEA/CUA | Gastric cancer and Gastroesophageal junction cancer | Nivolumab + Chemotherapy | Health system or payer perspective | Lifetime (10 years) | Absent | Not cited |
| Kuznik et al., 2022^193^ | US | CEA/CUA | Non-small cell lung cancer | Cemiplimab | Health system or payer perspective | Lifetime (30 years) | Present | Not cited |
| Leung et al., 2022^194^ | Taiwan | CUA | Non-small cell lung cancer | Atezolizumab; Nivolumab; Pembrolizumab | Health system or payer perspective | Lifetime (10 years) | Absent | Not cited |
| Li et al., 2022^195^ | China | CEA/CUA | Hepatocellular carcinoma | Atezolizumab + Bevacizumab; Sintilimab + Bevacizumab biosimilar | Health system or payer perspective | 15 years | Absent | Not cited |
| Li et al., 2022^196^ | China | CUA | Esophageal cancer | Camrelizumab | Health system or payer perspective | 5 years | Absent | Cited |
| Li et al., 2022^197^ | China | CUA | Non-small cell lung cancer | Sugemalimab + Chemotherapy | Health system or payer perspective | 10 years | Absent | Not cited |
| Lin et al., 2022^198^ | China | CUA | Esophageal cancer | Nivolumab | Health system or payer perspective | 36 months | Absent | Cited |
| Lin et al., 2022^199^ | US | CUA | Non-small cell lung cancer | Atezolizumab | Health system or payer perspective | Lifetime | Absent | Not cited |
| Liu et al., 2022^200^ | China | CUA | Non-small cell lung cancer | Sintilimab + Chemotherapy | Health system or payer perspective | 10 years | Absent | Not cited |
| Liu et al., 2022^201^ | China | CEA/CUA | Small cell lung cancer | Durvalumab + Etoposide + Platinum | Health system or payer perspective | 10 years | Absent | Not cited |
| Liu et al., 2022^202^ | US | CEA/CUA | Endometrial carcinoma | Pembrolizumab + Lenvatinib | Health system or payer perspective | 7 years | Absent | Not cited |
| Liu et al., 2022^203^ | China | CEA/CUA | Hepatocellular carcinoma | Camrelizumab + Rivoceranib; Sintilimab + Bevacizumab biosimilar; Tislelizumab | Health system or payer perspective | 15 years | Absent | Cited |
| Liu et al., 2022^204^ | China and US | CEA/CUA | Urothelial carcinoma | Atezolizumab + Chemotherapy | Health care sector perspective | 10 years | Absent | Not cited |
| Liu et al., 2022^205^ | US | CBA/CEA/CUA | Renal cell carcinoma | Nivolumab + Cabozantinib | Health system or payer perspective | 10 years | Absent | Not cited |
| Luo et al., 2022^206^ | China | CUA | Non-small cell lung cancer | Tislelizumab + Pemetrexed-platinum chemotherapy | Health system or payer perspective | 20 years | Absent | Not cited |
| Mo et al., 2022^207^ | Japan | CUA | Non-small cell lung cancer | Ipilimumab + Nivolumab | Health system or payer perspective | 20 years | Absent | Not cited |
| Peng et al., 2022^208^ | China | CEA/CUA | Hepatocellular carcinoma | Sintilimab + Bevacizumab biosimilar | Health system or payer perspective | Lifetime | Absent | Not cited |
| Polyzoi et al., 2022^209^ | US | CEA/CUA | Non-small cell lung cancer | Ipilimumab + Nivolumab + Platinum-doublet chemotherapy | Health system or payer perspective | 25 years | Present | Not cited |
| Qu et al., 2022^210^ | US | CBA/CEA/CUA | Esophageal cancer | Pembrolizumab + Cisplatin + 5-Fluorouracil | Health system or payer perspective | Lifetime (37.6 years) | Present | Not cited |
| Rui et al., 2022^211^ | China | CEA/CUA | Non-small cell lung cancer | Sintilimab + Chemotherapy | Health system or payer perspective | Lifetime | Present | Not cited |
| She et al., 2022^212^ | US | CEA/CUA | Head and neck cancer | Pembrolizumab; Pembrolizumab + Cetuximab | Health system or payer perspective | Lifetime | Absent | Not cited |
| Shen et al., 2022^213^ | China | CEA/CUA | Esophageal cancer | Sintilimab + Chemotherapy | Health system or payer perspective | Lifetime | Absent | Not cited |
| Shi et al., 2022^214^ | China | CUA | Esophageal cancer | Tislelizumab | Health system or payer perspective | 10 years | Absent | Not cited |
| Shi et al., 2022^215^ | US | CEA/CUA | Cervical cancer | Pembrolizumab | Health system or payer perspective | 30 years | Absent | Cited |
| Shu et al., 2022^216^ | China | CUA | Gastric cancer, Gastroesophageal junction cancer, and Esophageal adenocarcinoma | Nivolumab + Chemotherapy | Health system or payer perspective | 5 years | Absent | Not cited |
| Song et al., 2022^217^ | China | CUA | Esophageal cancer | Pembrolizumab | Health system or payer perspective | 10 years | Absent | Not cited |
| Tang et al., 2022^218^ | China | CEA/CUA | Melanoma | Pembrolizumab | Health system or payer perspective | 20 years | Present | Not cited |
| Tian et al., 2022^219^ | China | CEA/CUA | Head and neck cancer (Nasopharyngeal carcinoma) | Camrelizumab + Cisplatin + Gemcitabine; Toripalimab + Cisplatin + Gemcitabine | Health system or payer perspective | Lifetime | Absent | Not cited |
| Tong et al., 2022^220^ | China | CUA | Small cell lung cancer | Durvalumab + Etoposide + Platinum | Health system or payer perspective | Lifetime | Absent | Not cited |
| Wahler et al., 2022^221^ | Germany | CEA/CUA | Melanoma | Nivolumab | Health system or payer perspective | Lifetime | Present | Not cited |
| Wang et al., 2022^222^ | China | CEA/CUA | Renal cell carcinoma | Nivolumab + Cabozantinib | Health system or payer perspective | 20 years | Absent | Not cited |
| Wang et al., 2022^223^ | China | CEA/CUA | Non-small cell lung cancer | Sugemalimab + Platinum-based chemotherapy | Health system or payer perspective | 10 years | Absent | Not cited |
| Wang et al., 2022^224^ | China | CEA/CUA | Renal cell carcinoma | Pembrolizumab + Lenvatinib | Health system or payer perspective | 5 years | Absent | Not cited |
| Wu et al., 2022^225^ | China | CEA/CUA | Esophageal cancer | Pembrolizumab + Chemotherapy | Health system or payer perspective | Lifetime | Absent | Not cited |
| Xie et al., 2022^226^ | China and US | CUA | Urothelial carcinoma | Avelumab + Best supportive care | Health system or payer perspective | 10 years | Absent | Not cited |
| Xie et al., 2022^227^ | US | CUA | Esophageal cancer | Pembrolizumab | Health system or payer perspective | 10 years | Absent | Not cited |
| Yang et al., 2022^228^ | US | CEA/CUA | Malignant pleural mesothelioma | Ipilimumab + Nivolumab | Health system or payer perspective | 10 years | Absent | Not cited |
| Ye et al., 2022^229^ | China | CEA/CUA | Esophageal cancer | Sintilimab + Chemotherapy | Health system or payer perspective | 10 years | Absent | Cited |
| Ye et al., 2022^230^ | US | CEA/CUA | Malignant pleural mesothelioma | Ipilimumab + Nivolumab | Health system or payer perspective | 10 years | Absent | Cited |
| Yeh et al., 2022^231^ | Not reported | CUA | Head and neck cancer | Pembrolizumab | Health system or payer perspective | Not reported | Absent | Not cited |
| You et al., 2022^232^ | China | CUA | Esophageal cancer | Sintilimab + Chemotherapy | Health system or payer perspective | 6 years | Absent | Not cited |
| Zhan et al., 2022^233^ | China | CUA | Esophageal cancer | Pembrolizumab | Societal perspective | 5 years | Absent | Not cited |
| Zhang et al., 2022^234^ | China and US | CUA | Urothelial carcinoma | Atezolizumab + Chemotherapy | Health system or payer perspective | 15 years | Absent | Not cited |
| Zhang et al., 2022^235^ | US | CBA/CEA/CUA | Non-small cell lung cancer | Cemiplimab | American perspective | 20 years | Absent | Not cited |
| Zhao et al., 2022^236^ | China | CEA/CUA | Hepatocellular carcinoma | Atezolizumab + Bevacizumab; Sintilimab + Bevacizumab | Health system or payer perspective | Lifetime (10 years) | Absent | Not cited |
| Zheng et al., 2022^237^ | China | CEA/CUA | Esophageal cancer | Pembrolizumab + Chemotherapy | Health system or payer perspective | 10 years | Absent | Not cited |
| Zhou et al., 2022^238^ | China | CEA/CUA | Hepatocellular carcinoma | Sintilimab + Bevacizumab biosimilar | Health system or payer perspective | Lifetime | Absent | Cited |
| Zhou et al., 2022^239^ | China | CUA | Non-small cell lung cancer | Tislelizumab | Health system or payer perspective | 30 years | Absent | Not cited |
| Zhou et al., 2022^240^ | China | CEA/CUA | Hepatocellular carcinoma | Sintilimab + Bevacizumab | Health system or payer perspective | Lifetime | Absent | Not cited |
| Zhu et al., 2022^241^ | China | CEA/CUA | Small cell lung cancer | Serplulimab + Chemotherapy | Health system or payer perspective | 10 years | Absent | Cited |
| Zhu et al., 2022^242^ | China | CEA/CUA | Head and neck cancer (Nasopharyngeal carcinoma) | Toripalimab + Cisplatin + Gemcitabine | Health system or payer perspective | 10 years | Absent | Cited |
| Zhu et al., 2022^243^ | China and US | CEA/CUA | Esophageal cancer | Pembrolizumab + Cisplatin + 5-Fluorouracil | Health system or payer perspective | 7 years | Absent | Cited |
| Cheng et al., 2023^244^ | China | CUA | Non-small cell lung cancer | Sintilimab | Health system or payer perspective | 20 years | Absent | Not cited |
| Chu et al., 2023^245^ | Ireland | CEA/CUA | Non-small cell lung cancer | Pembrolizumab | Health system or payer perspective | 20 years | Absent | Not cited |
| Dioun et al., 2023^246^ | US | CUA | Endometrial carcinoma | Dostarlimab | Societal perspective | 2 years | Present | Not cited |
| Fei et al., 2023^247^ | China | CEA/CUA | Small cell lung cancer | Nivolumab; Pembrolizumab | Health system or payer perspective | 4 years | Absent | Not cited |
| Lin et al., 2023^248^ | US | CEA/CUA | Urothelial carcinoma | Avelumab + Best supportive care | Health system or payer perspective | Lifetime | Absent | Not cited |
| Sharma et al., 2023^249^ | US | CUA | Renal cell carcinoma | Pembrolizumab | Health system or payer perspective | 5 years; 15 years | Absent | Not cited |
| Shu et al., 2023^250^ | China | CBA/CUA | Non-small cell lung cancer | Ipilimumab + Nivolumab | Health system or payer perspective | 10 years | Absent | Not cited |
| Ye et al., 2023^251^ | US | CEA/CUA | Esophageal cancer | Pembrolizumab + Chemotherapy | Health system or payer perspective | 10 years | Absent | Cited |
| Zhu et al., 2023^252^ | US | CEA/CUA | Renal cell carcinoma | Pembrolizumab + Lenvatinib | Health system or payer perspective | 20 years | Absent | Not cited |

CBA indicates cost-benefit analysis; CEA, cost-effectiveness analysis; CHEERS, Consolidated Health Economic Evaluation Reporting Standards; CMA, cost-minimization analysis; CUA, cost-utility analysis; UK, the United Kingdom; US, the United States.
